# Effectiveness of Gastric Cancer Endoscopic Screening in Intermediate-Risk Countries – a Systematic Review and Meta-Analysis

**DOI:** 10.1101/2025.04.06.25325320

**Authors:** B. Mourato, N. Pratas, A Branco, R. Chança, I. Fronteira, R. Dinis, M. Areia

**Affiliations:** NOVA National School of Public Health, Public Health Research Centre, Comprehensive Health Research Center, CHRC, LA-REAL, CCAL, NOVA University Lisbon, Lisbon, Portugal, Lisbon, Portugal; Unidade Local de Saúde do Alto Alentejo, Hospital Doutor José Maria Grande, Portalegre, Portugal; Unidade Local de Saúde do Alto Alentejo; Divisão de Avaliação de Tecnologias em Saúde, Instituto Nacional de Câncer; NOVA National School of Public Health, Public Health Research Centre, Comprehensive Health Research Center, CHRC, LA-REAL, CCAL, NOVA University Lisbon, Lisbon, Portugal, Lisbon; Unidade Local de Saúde do Alentejo Central, Hospital do Espírito Santo de Évora, Évora, Portugal; Comprehensive Health Research Center, CHRC, Lisbon, Portugal; Instituto Português de Oncologia de Coimbra, RISE@CI-IPO (Health Research Network), Portuguese Oncology Institute of Porto (IPO Porto)

**Author notes:** Corresponding author: Beatriz Mourato Affiliation: NOVA National School of Public Health, Public Health Research Centre, Comprehensive Health Research Center, CHRC, LA-REAL, CCAL, NOVA University Lisbon, Lisbon, Portugal, Lisbon, Portugal; Unidade Local de Saúde do Alto Alentejo, Hospital Doutor José Maria Grande, Portalegre, Portugal; Address: Hospital Doutor José Maria Grande, Avenida de Santo António, 7300- 853, Portalegre; Portugal, Contacts. Contacts:; Address: Hospital Doutor José Maria Grande, Avenida de Santo António, 7300-853, Portalegre Contributions: conceptualization, designing, investigation, formal analysis and writing the manuscript (B. Mourato). Contacts:; Address: Hospital Doutor José Maria Grande, Avenida de Santo António, 7300- 853, Portalegre; Contributions: provided assistance with protocol design, investigation, and writing (N. Pratas). Contacts:; Address: Hospital Doutor José Maria Grande, Avenida de Santo António, 7300- 853, Portalegre; Contributions: for investigation, and reviewing the writing of the manuscript;4 (A Branco). Contacts; Address: Contributions: designing the search expression in the various databases (R. Chança). Contacts:; Address: Avenida Padre Cruz, 1600-560 Lisboa; Contributions: protocol design, systematic review process supervision, and manuscript review (I. Fronteira). Contacts:; Address: Largo do Sr. Da Probreza, 7000-811 Évora; Contributions: revision of the manuscript. Contacts:; Address: Avenida Bissaya Barreto 98, 3000-075 Coimbra. Contributions: protocol design, systematic review process supervision, and manuscript review.

**Keywords:** Stomach Neoplasm, Early Detection of Cancer, Mass Screening Endoscopy, Risk Factors, diagnosis, survival, meta-analysis, Gastrointestinal Neoplasms, Public Health

## Abstract

**Background:** Gastric cancer remains a major cause of cancer mortality worldwide, including in intermediate-risk countries. While endoscopic screening has proven effective in high-risk populations, its impact and economic value in intermediate-risk settings remain uncertain.

**Objective:** This systematic review and meta-analysis aimed to evaluate the effectiveness and cost-effectiveness of endoscopic screening for gastric cancer in these countries.

**Design:** A systematic review and meta-analysis was conducted following a comprehensive search in Medline, Scopus, Embase, and Web of Science, covering studies published up to 30 September 2024. A random-effects meta-analysis was performed for effectiveness studies, and economic evaluations were synthesised narratively. The study **registration** number at PROSPERO is CRD42024502174.

**Results:** Thirty-two studies met inclusion criteria—24 on screening effectiveness and eight on cost-effectiveness. Across the 24 effectiveness studies, 404,159 individuals underwent upper endoscopic screening, which significantly increased detection of precancerous lesions (pooled effect size: 28%, p < 0.001) and early-stage gastric cancer among neoplasms (73.6%, p < 0.001). Screening was also associated with a 26.1% reduction in gastric cancer mortality and improved five-year survival (63.7% to 85.0%). Economic analyses suggested endoscopic screening is cost-effective in intermediate-risk settings, particularly when combined with colorectal cancer screening.

**Conclusions:** Endoscopic screening improves early detection and survival in intermediate-risk countries. Cost-effectiveness studies support its feasibility, especially when integrated with colorectal cancer screening or risk-stratified strategies.

**KEY MESSAGES:** *What is already known on this topic:* Endoscopic screening reduces gastric cancer mortality in high-risk countries. Evidence in intermediate-risk settings remains limited.

*What this study adds:* This review shows that endoscopic screening improves early detection and gastric cancer prognosis in intermediate-risk countries and is cost-effective in several models.

*How this study might affect research, practice or policy:* Findings support implementing screening where cost-effectiveness is demonstrated, aligning with ESGE MAPS III recommendations.

## INTRODUCTION

Gastric cancer (GC) is the fifth most common and fifth deadliest cancer worldwide, with significant geographical variation.^1^ High-risk countries such as Japan, Mongolia, and South Korea report age-standardised incidence rates ≥20 per 100,000 person-years, while intermediate-risk countries, including China, Iran, Peru, and Portugal, present age-standardised incidence rates between 10 and 20.^1^ Although global incidence is declining, there is a concerning rise in GC among younger individuals and in proximal or diffuse tumour types, which are often associated with poorer outcome.^2,3^

GC is typically asymptomatic in early stages, resulting in delayed diagnosis and poor prognosis.^4,5^ Five-year survival is below 40% in advanced disease^6^ but exceeds 90% when diagnosed early, allowing for minimally invasive treatments such as endoscopic resection.^7^ However, symptom-based detection is largely ineffective for early diagnosis.

Oesophagogastroduodenoscopy (EGD) is the gold standard for GC screening.^8,9^ In high-risk countries, such as Japan and South Korea, national EGD-based programmes have demonstrated significant reductions in mortality.^10,11^ China has implemented structured screening initiatives in both rural and urban populations.^5^ Conversely, most intermediate-risk countries lack national screening strategies despite emerging evidence supporting the value of EGD.

Recent studies suggest that EGD screening, particularly when combined with colorectal cancer screening, may be cost-effective in intermediate-risk settings.^12,13^ This position is supported by United European Gastroenterology and the European Society of Gastrointestinal Endoscopy.^14,15^ Moreover, the recently published Management of epithelial precancerous conditions and early neoplasia of the stomach III guidelines, issued by the European Society of Gastrointestinal Endoscopy in 2025, reinforce these recommendations by stating that endoscopic screening may be considered in intermediate-risk countries, if cost-effectiveness has been demonstrated and resources are available.^16^

Despite increasing interest, there is no comprehensive synthesis of the effectiveness and cost-effectiveness of EGD screening in intermediate-risk populations. This systematic review and meta-analysis aim to address this gap by evaluating EGD’s impact on detection of precancerous lesions, GC diagnosis (overall and early-stage), survival, and mortality. Additionally, we assess the economic viability of EGD screening based on programme costs and incremental cost-effectiveness ratios (ICERs).

## METHODS

This review adhered to the Preferred Reporting Items for Systematic Reviews and Meta-Analyses (PRISMA) guidelines and defined eligibility using the Population, Intervention, Comparison, Outcomes, and Study (PICOS) framework (table 1). The protocol was registered with PROSPERO (CRD42024502174) and published with detailed information.^17^

**Table 1.** Population, Intervention, Comparison, Outcome and Study Design framework for this systematic review and meta-analysis.

| Question: What is the effectiveness of endoscopic screening for gastric cancer in intermediate-risk countries? |  |
| --- | --- |
| Population | Asymptomatic population of intermediated-risk countries (countries with incidence age-standardized rate 10-20 per 100 000 person/years: Tajikistan, Iran, Azerbaijan, Kyrgyzstan, Bhutan, Belarus, Peru, Mali, Chile, Costa Rica, Democratic People Republic of Korea, China, Kazakhstan, Russian Federation, Vietnam, Estonia, Colombia, Portugal, Ecuador, Albania, Guadeloupe (France), Guatemala, Latvia, Armenia, Turkmenistan, Myanmar, Samoa, Turkey, Lithuania, Lao People's Democratic Republic, Sao Tome and Principe, Afghanistan, Martinique (France), Brunei Darussalam, Zimbabwe, Uzbekistan), between 40 and 80 years of age, without diagnostic of GC or precancerous conditions. |
| Intervention | Endoscopic screening for gastric cancer. |
| Comparison | No screening for gastric cancer. |
| Outcome | The effectiveness of endoscopic screening of gastric cancer was defined as: detection rate of precancerous lesions; detection rate of gastric cancer; detection rate of early gastric cancer; stage at diagnosis; mortality rate of gastric cancer of screened vs non-screened patients; 5-year survival rate of gastric cancer screened; costs of screening program. |
| Studies Designs | Randomized controlled trials, non-randomized controlled trials, cohort studies, case-control studies, cross-sectional studies and cost-effectiveness studies. |

A comprehensive search was conducted in Medline, Scopus, Embase, and Web of Science, supplemented by grey literature and reference lists. Search strategy adhered to Peer Review of Electronic Search Strategies guidelines^18^ and was tailored to each database (Supplementary File 1 - Search strategy). We included randomised controlled trials, non-randomised controlled trials, cohort studies, case-control studies, cross-sectional studies, other observational studies, cost-effectiveness, cost-utility, and decision-analytic models published as free full-text articles in English, Portuguese, or Spanish, up to 30 September 2024.

Two reviewers independently screened title, abstracts and full texts with disagreements resolved by discussion and consultation with additional reviewers. When multiple publications described the same cohort, the most comprehensive or recent was included.

Data extraction was performed independently by three reviewers using a predefined form (Supplementary File 2 – variables form). Studies were classified into effectiveness or economic analyses.

The risk of bias was assessed with appropriate tools: Risk of Bias 2 for randomised controlled trials, Risk of Bias for non-randomised studies of interventions, the Newcastle-Ottawa Quality Assessment Scale^19^ for case-control and cohort studies, the National Heart, Lung, and Blood Institute study quality assessment tools^20^ for cross-sectional studies, and the Consensus on Health Economic Criteria list^21^ for cost-effectiveness studies.

A meta-analysis of effectiveness outcomes was performed using Jamovi (v2·4·8), applying a random-effects model with Restricted Maximum Likelihood estimation to derive pooled effect sizes for five primary outcomes: detection of precancerous conditions, detection of gastric cancer (positive lesions), early-stage gastric cancer (EGC) detection, five-year survival, and gastric cancer-related mortality. “Positive lesions” were defined as high-grade intraepithelial neoplasia, carcinoma in situ, suspicious for invasive carcinoma, and invasive neoplasia (Vienna Classification categories 4 and 5).^22^ Statistical significance was assessed using a Z-test^23^, with a p-value threshold of <0.05 considered statistically significant.^24^ Confidence intervals (CI) of 95% were reported for all estimates. Heterogeneity was assessed using Cochran’s Q test, and quantified by I² and Tau².^23^ The I² statistic represents the proportion of total variability attributable to heterogeneity, categorised as low (0-25%), moderate (25-50%), high (50-75%), or very high (>75%), with high values indicating considerable variability that may require further analysis.^25^ The Tau² statistic estimates the absolute between-study variance, with higher values indicating greater variability in effect sizes across studies.^26^

A systematic review without meta-analysis was conducted for the economic evaluation, synthesising the cost-effectiveness outcomes of endoscopic screening in intermediate-risk countries.

## RESULTS

The search identified 1,615 records, with 969 remaining after duplicates were removed (Supplementary File 3 - All Studies Rayyan; Supplementary File 4 – Excluded Studies) (Figure 1).

**Figure 1.**
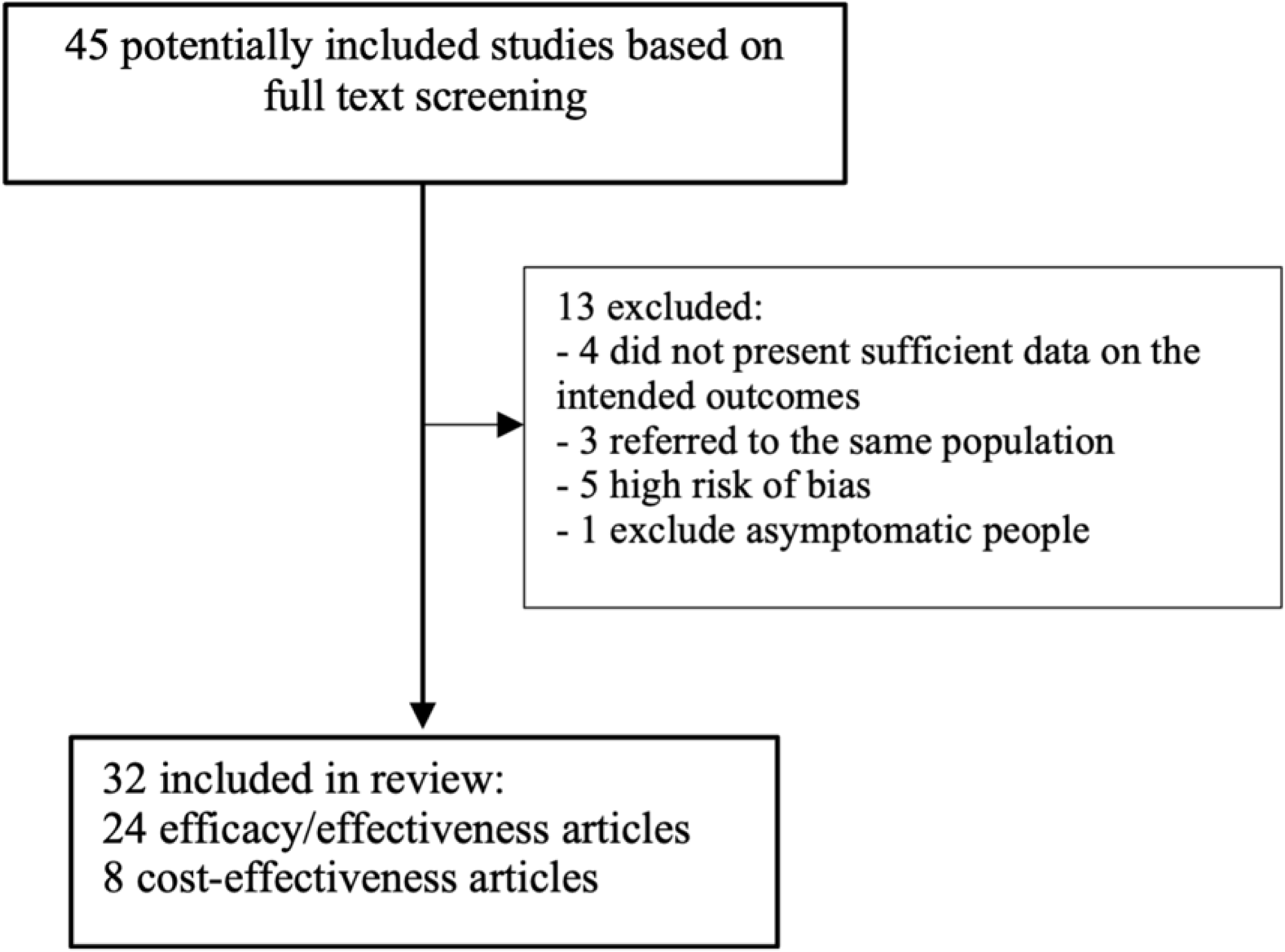
Flowchart of included studies according to the Preferred Reporting Items for Systematic Reviews and Meta-Analyses guidelines.

**Figure 2.**
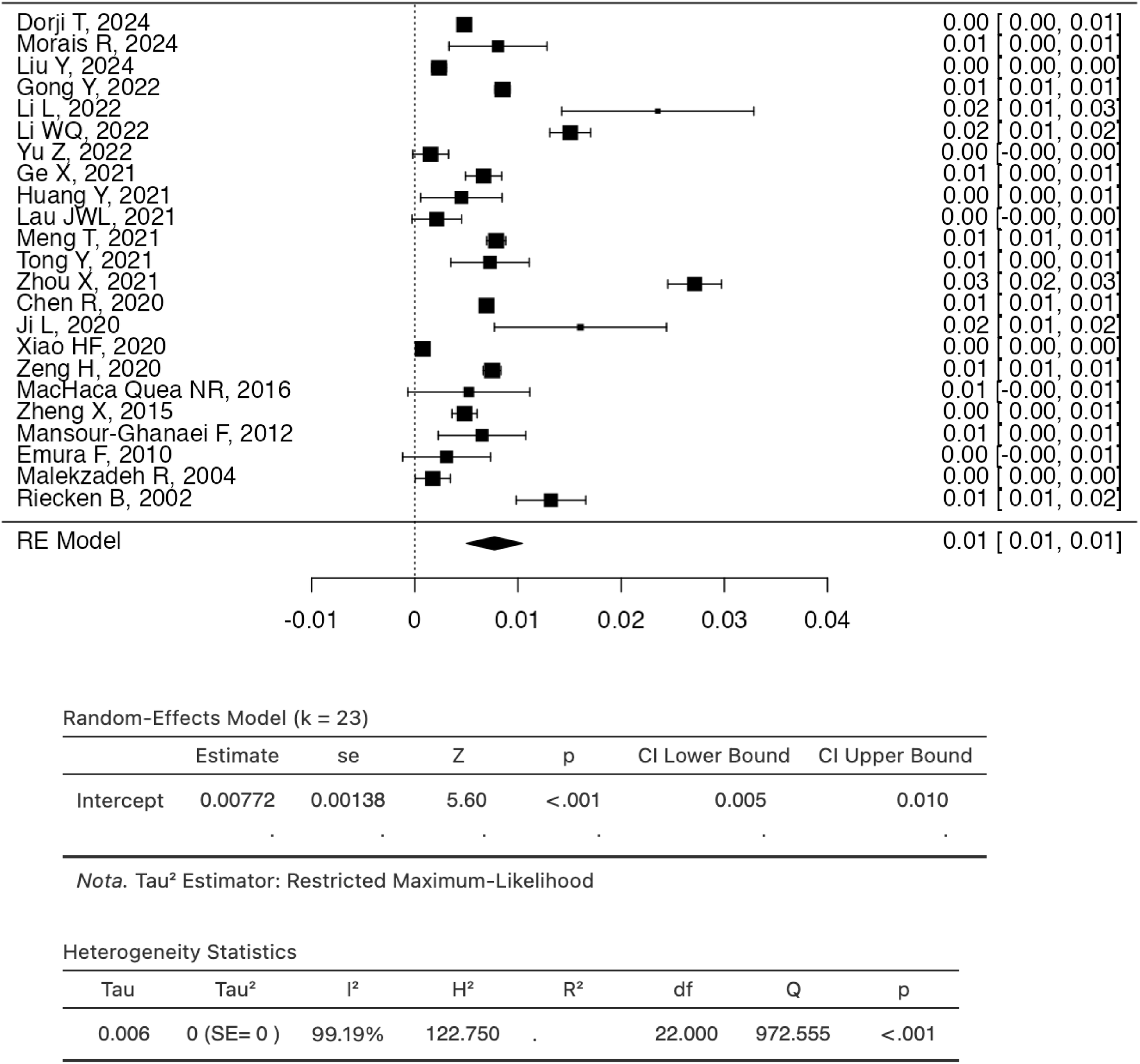
Flowchart of the final decision to include articles in this study.

Following title and abstract screening, 75 articles underwent full-text review (inter-reviewer agreement: 91.0%; kappa correlation coefficient: 0.82^27^), of which 43 met inclusion criteria (inter-reviewer agreement: 96%; kappa correlation coefficient: 0.92^28^).

An additional two eligible studies were identified through reference screening, resulting in 45 total included articles (Supplementary File 5 - Included Studies Dataset): 34 effectiveness studies; 11 cost-effectiveness studies. After risk of bias assessment, 24 studies were retained for the effectiveness analysis and 8 for cost-effectiveness (Figure2).

### Effectiveness Studies Analysis

Across the effectiveness studies, 404,159 individuals underwent EGD screening. The main characteristics of these studies are presented in Table 2.

**Table 2.**
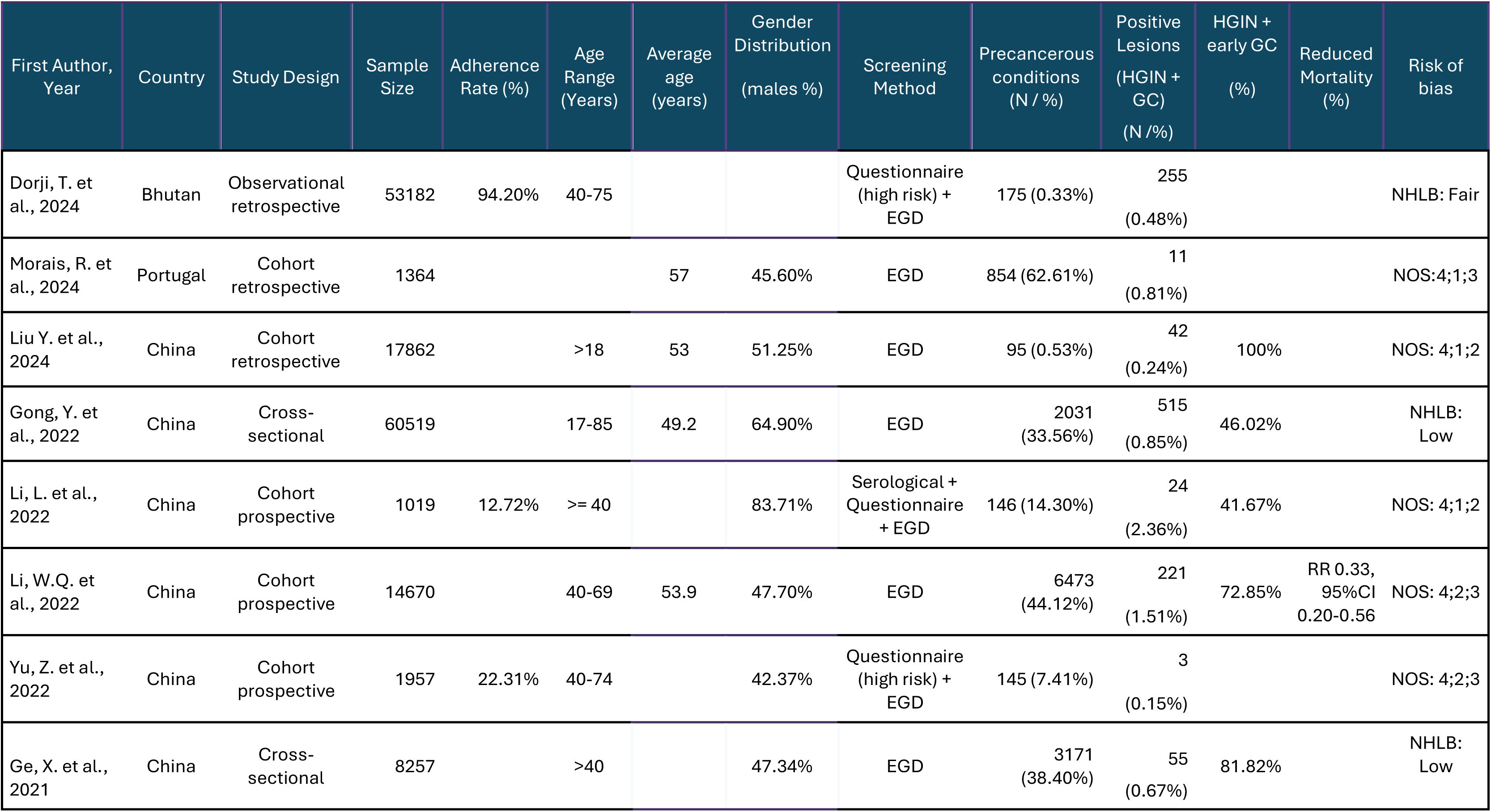
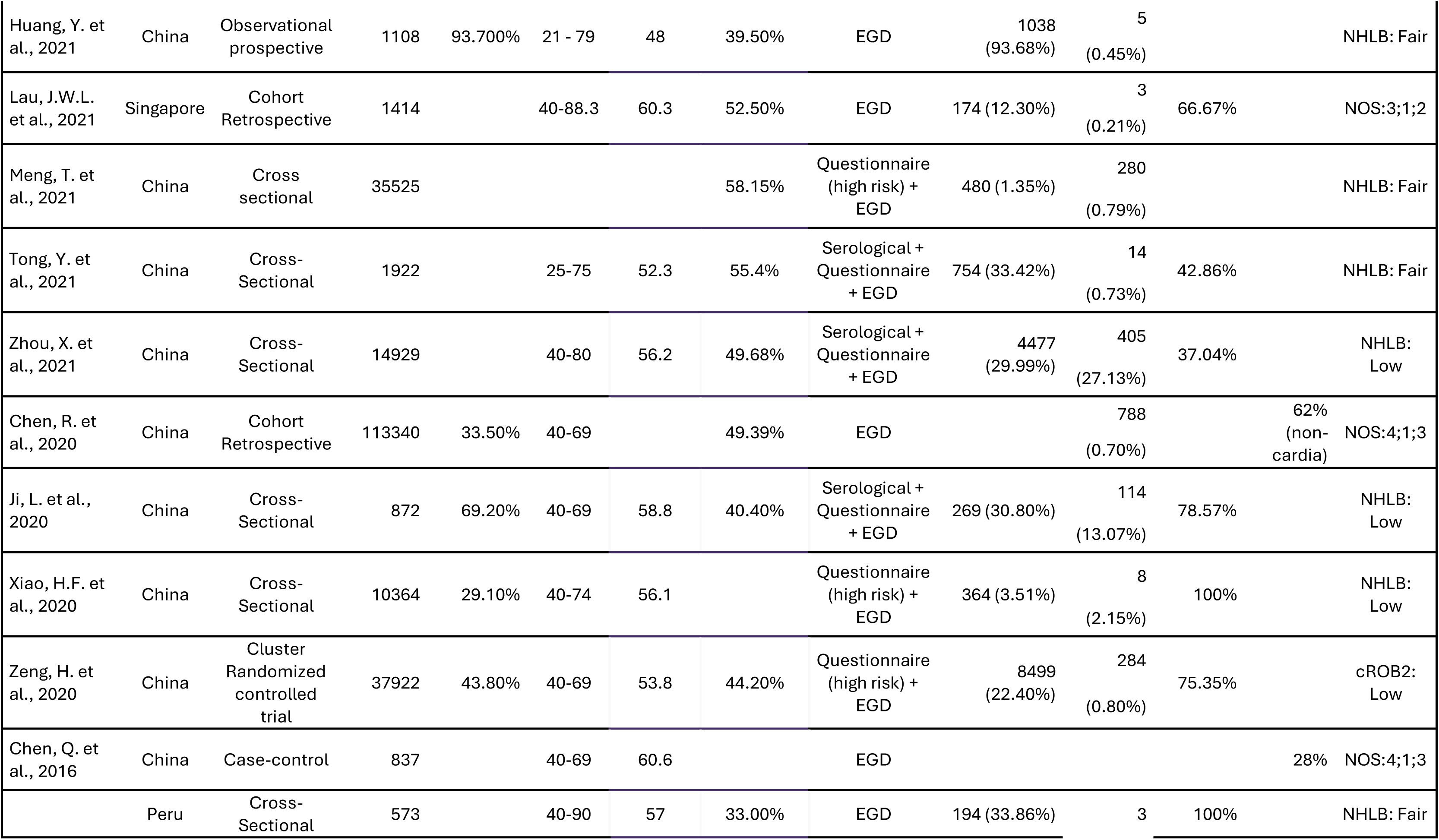
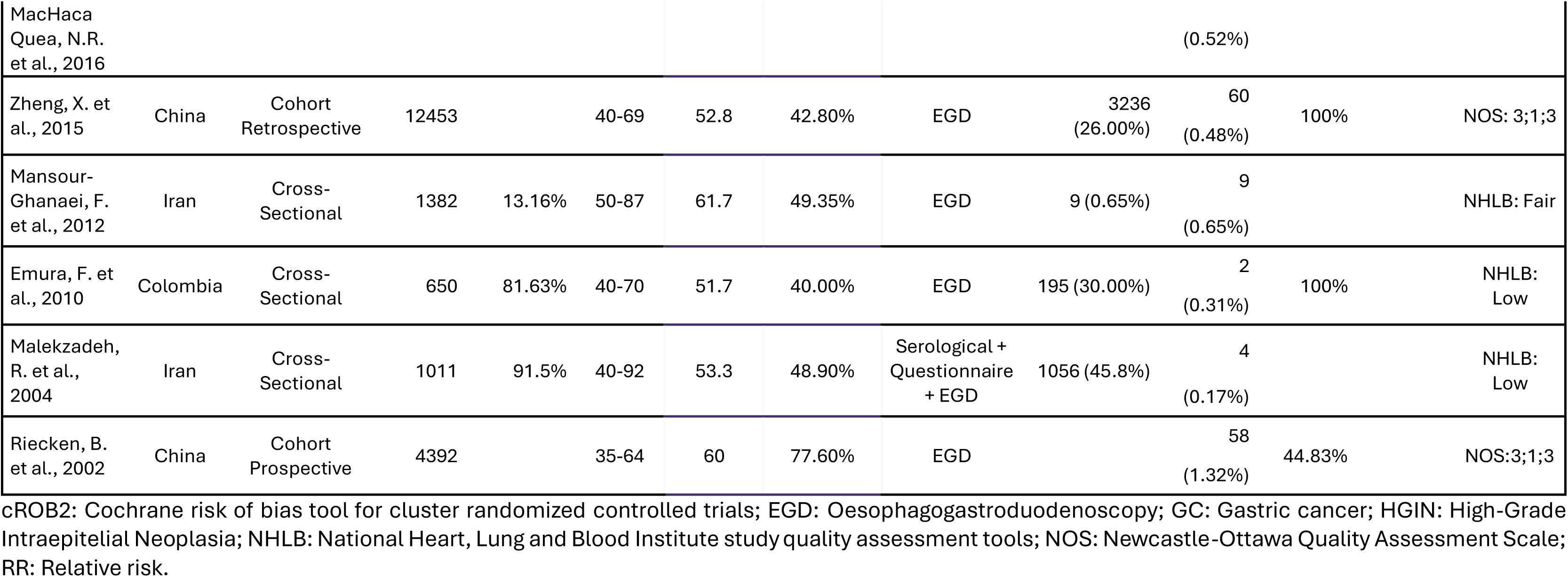
Characteristics of included effectiveness studies and a description of the main outcomes.

Most were conducted in China^29,30,30–45^, where two endoscopic screening programmes for GC were implemented. In addition, we found seven studies from other countries where endoscopic screening was not implemented, corresponding to some pilot programmes for this type of screening.^37,46–51^ Study designs included 1 cluster randomised controlled trial^29^, 1 case-control^32^, 4 prospective^34,39,40,42^ and 5 retrospective^33,44,50–52^ cohorts, 11 cross-sectional^31,36–38,41,46,47,49,53–55^, 1 prospective^43^, and 1 retrospective observational studies^48^.

Screening was mostly offered to individuals aged ≥40 years, with some studies incorporating risk stratification tools or serological tests prior to EGD.^29,35,36,38,39,41,42,45,47,48^ Adherence ranged from 12.7% to 94.2% (Table 2) and follow-up from 1 to 11.5 years. (Supplementary File 6 - Included studies outcomes dataset).

### Detection Rate Of Helicobacter Pylori And Precancerous Conditions

Helicobacter pylori prevalence, reported in 14 studies^34,36–39,41,43,46–51,54^, ranged from 13.1% to 89.2% (pooled effect size: 49.4% (p<0.001, Tau^2^=3.7%; I^2^=99.9%) (Supplementary File 7 - Helicobacter Pylori Meta-analysis).

Precancerous conditions (atrophic gastritis, intestinal metaplasia, and low-grade intraepithelial neoplasia) were reported in 22 studies^29–31,34–51,54^, with frequencies from 0.3% to 94.1% (pooled effect size: 28% (p<0.001; Tau^2^=6.60%; I^2^=100%) (Supplementary File 8 - Precancerous lesions Meta-analysis).

### Detection Rate Of Positive Lesions and EGC

Regarding positive cases (High-Grade Intraepithelial Neoplasia, EGC, and advanced GC), 23 studies^29–31,33–51,54^ reported prevalence rates ranging from 0.1% to 2.7% (pooled effect size: 0.8% (p<0·001; Tau^2^=0·0%; I²=99.2%; Supplementary File 9 - Positive lesions Meta-analysis) (Figures 3 and 4). The frequency of early-stage lesions (High-Grade Intraepithelial Neoplasia and EGC) among all detected neoplastic lesions was reported in 15 studies^43,45,46,48,50,51,54,55,57–60,62,66,67^, ranging from 37% to 100% (pooled effect size: 73.6% (p<0.001; Tau²=6.0%; I²=100%) (Supplementary File 10 - EGC Meta-analysis) (Figures 5 and 6).

**Figure 3.**
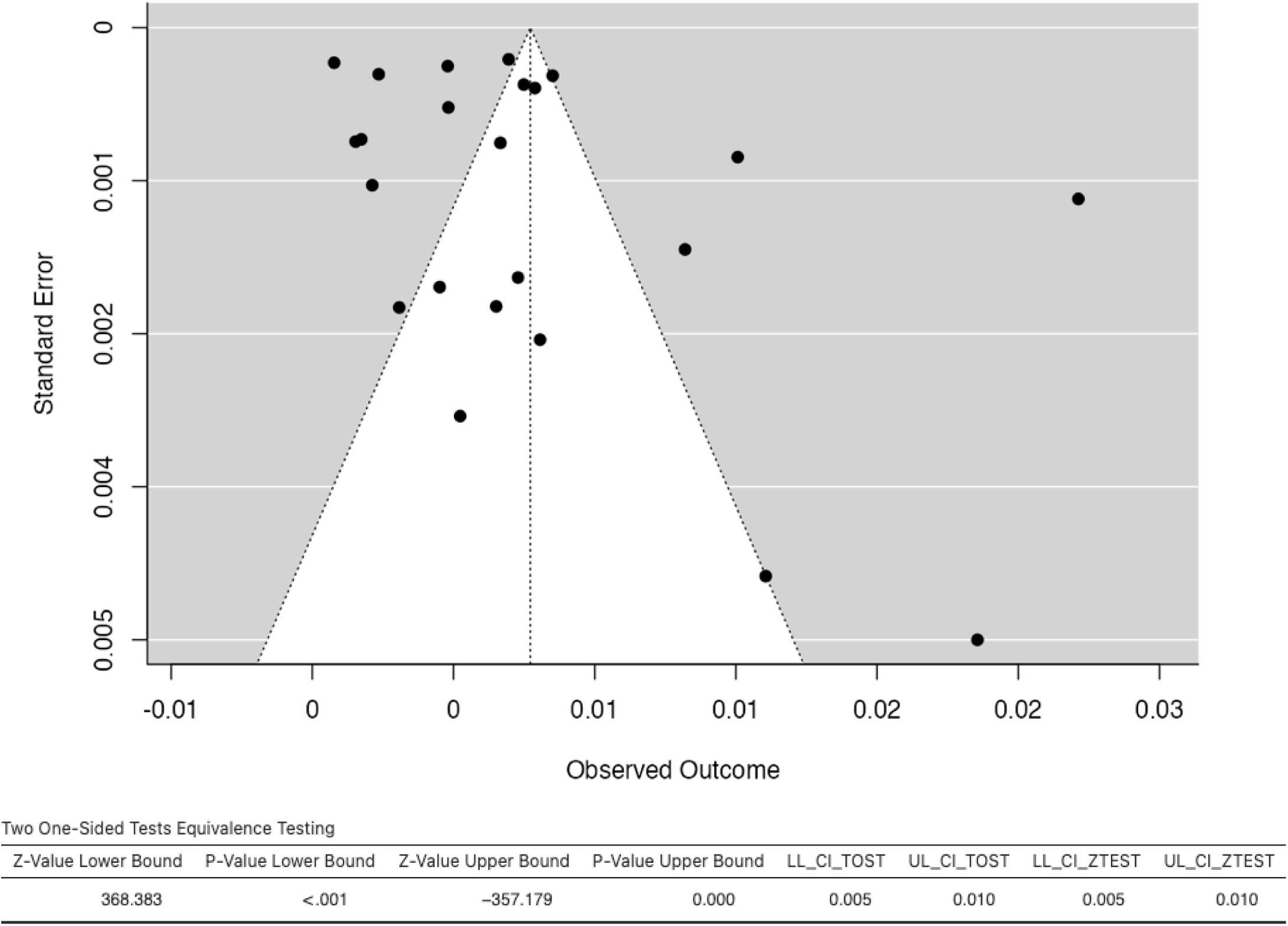
Forest Plot – Effect size for frequencies of positive lesions on endoscopic screening.

**Figure 4.**
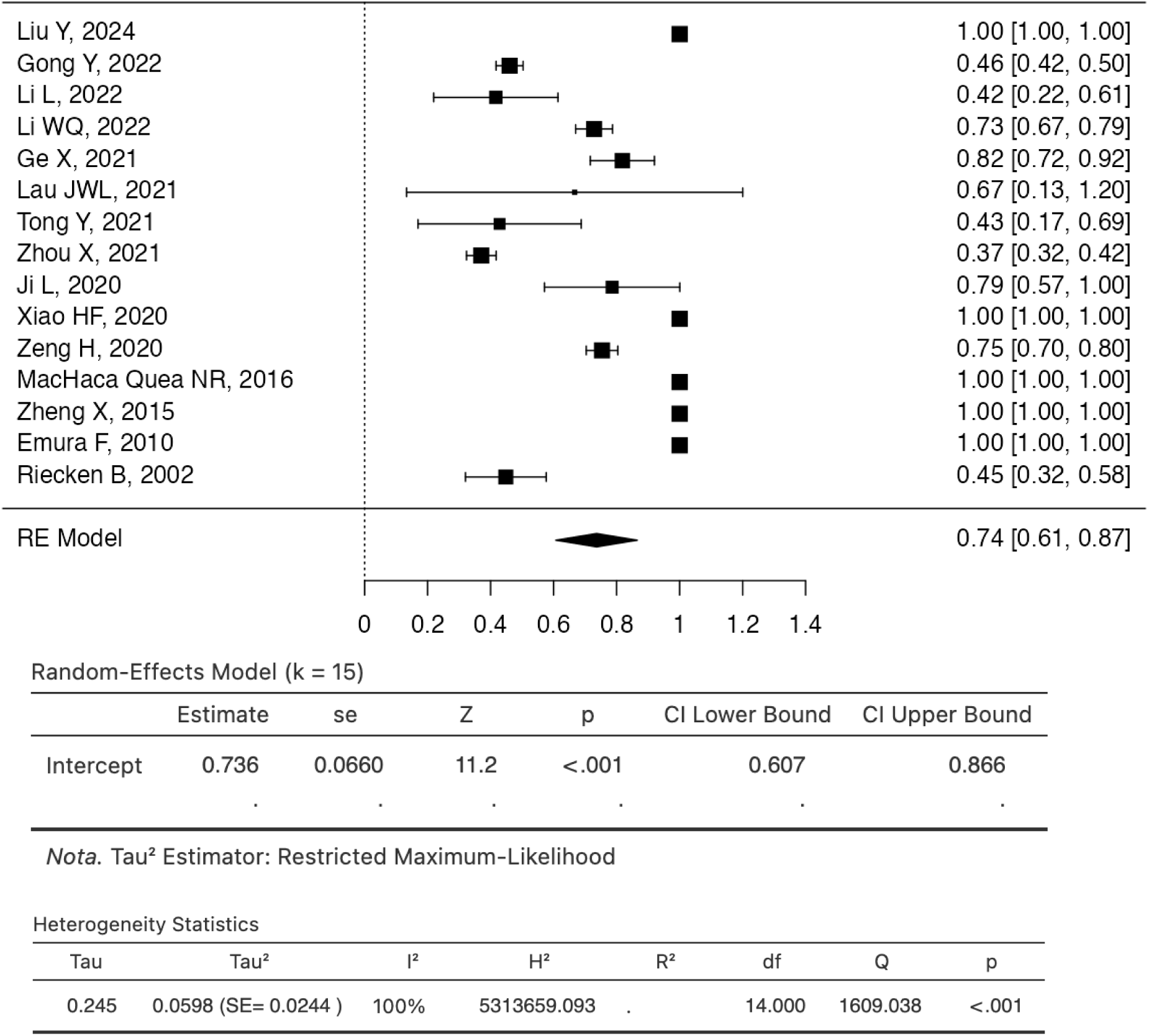
Funnel Plot for publication bias and heterogeneity in meta-analyse of positive lesions.

**Figure 5.**
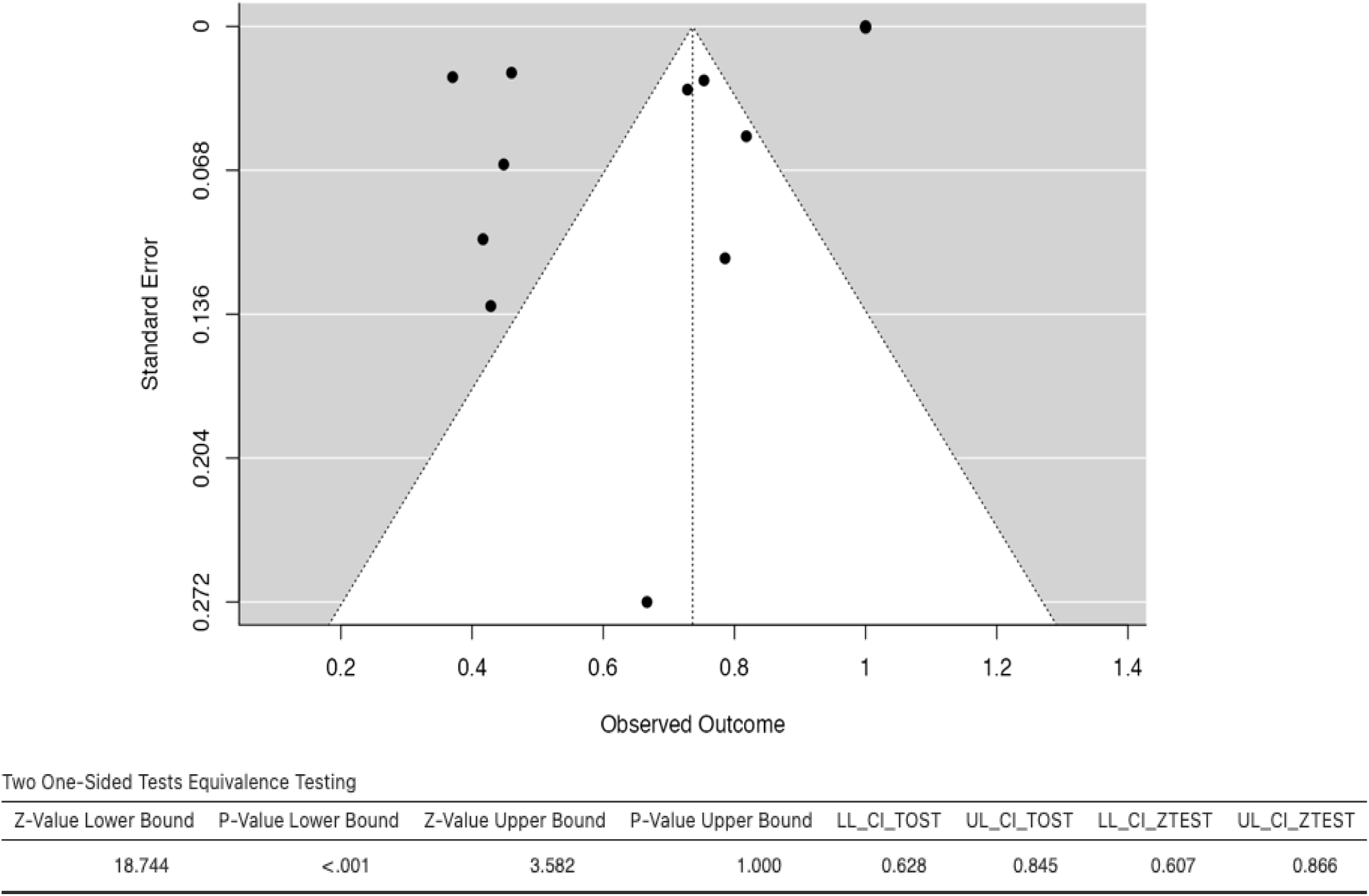
Forest Plot – Effect size for Frequencies of Hight-Grade intraepithelial Neoplasia and EGC among all neoplastic on endoscopic screening.

**Figure 6.**
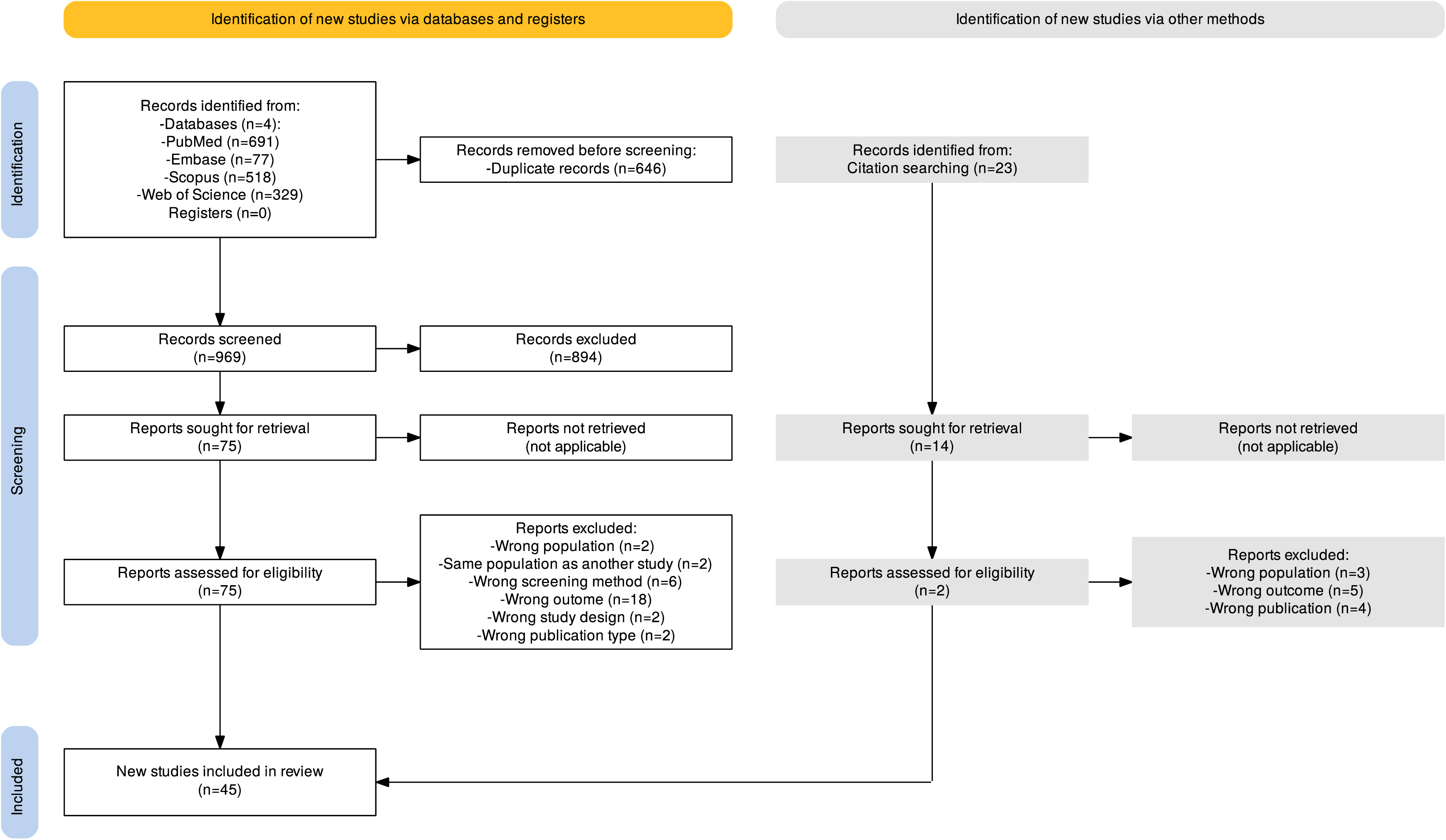
Funnel Plot for publication bias and heterogeneity in meta-analysis of High-Grade Intraepithelial Neoplasia and EGC among all neoplastic lesions on endoscopic screening.

### Effect Size Of Endoscopic Screening On Gc Mortality And Five-Year Survival

GC mortality was reported in five studies^30,32–34,39^; one^32^ lacked data on the total number of GC cases and was therefore excluded from the meta-analysis. The pooled mortality rate was 26.1% (p<0.001; I²=91%). Only three studies^32,33,39^ reported relative mortality risk reductions (28%–43%). Five-year survival was reported in three studies^30,34,40^, with rates from 63.7% to 85.0%.

### Cost-Effectiveness Studies Analysis

This review includes eight cost-effectiveness studies using microsimulation and Markov modelling techniques. Key characteristics are summarised in Table 3. Due to methodological heterogeneity, a narrative synthesis was performed.

**Table 3.**
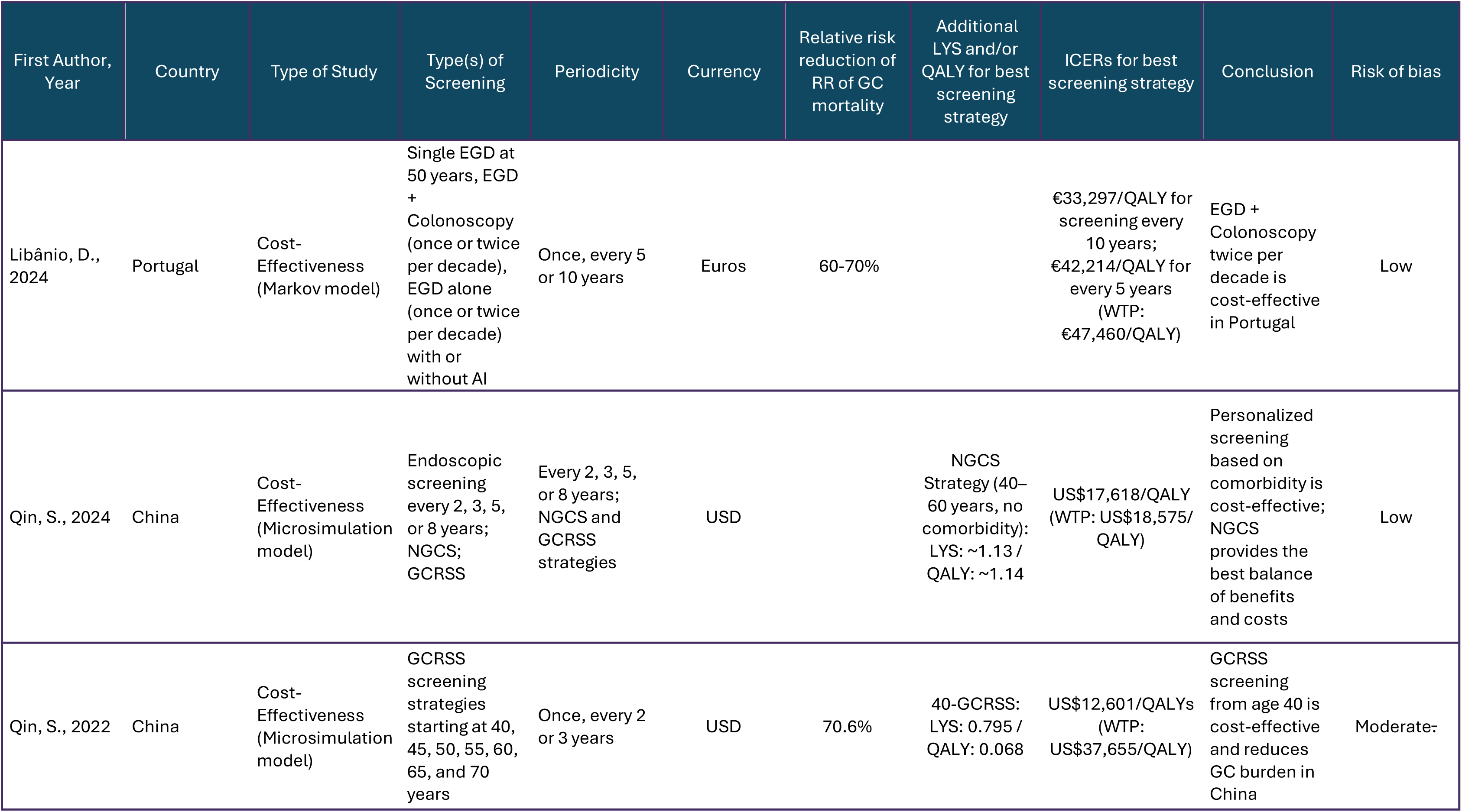
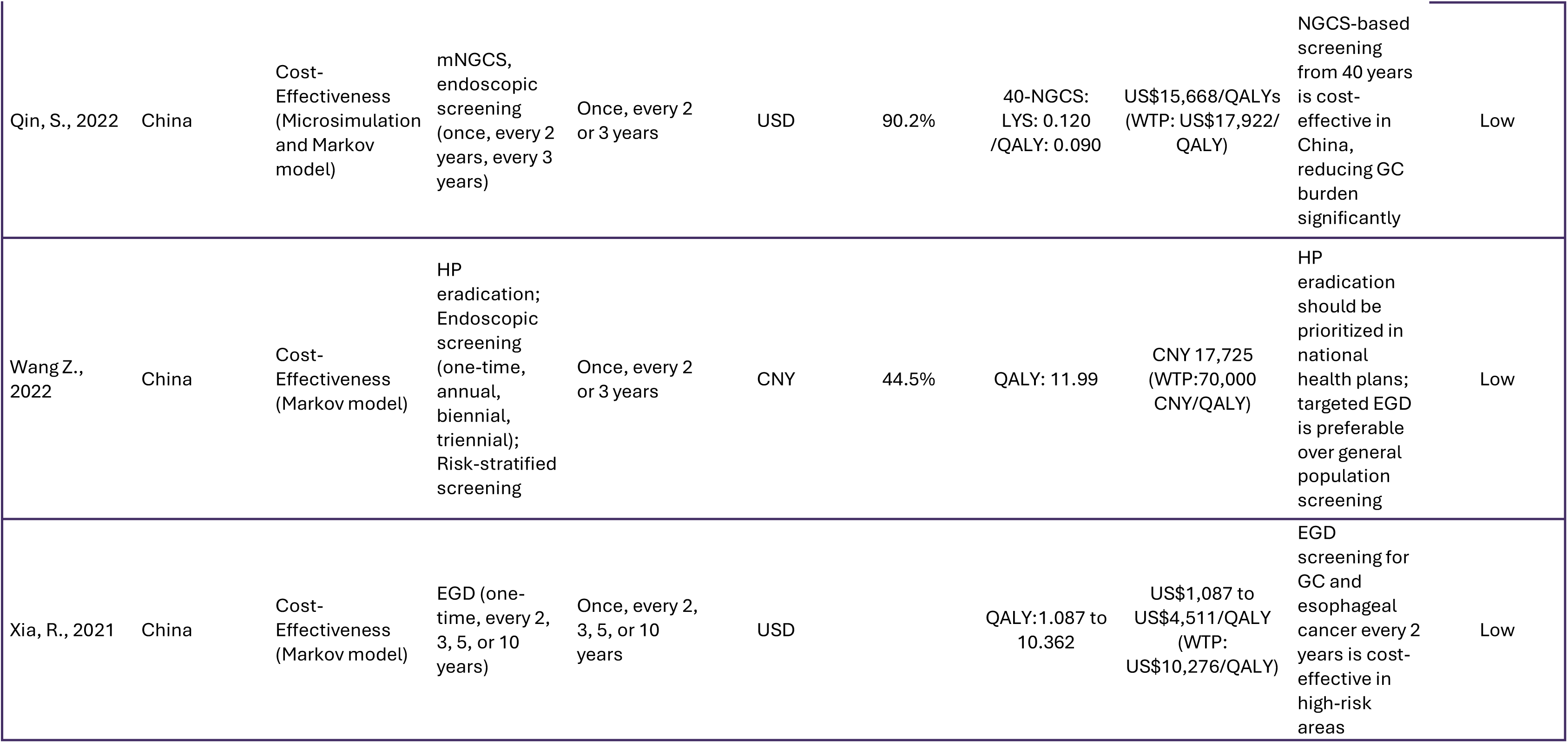
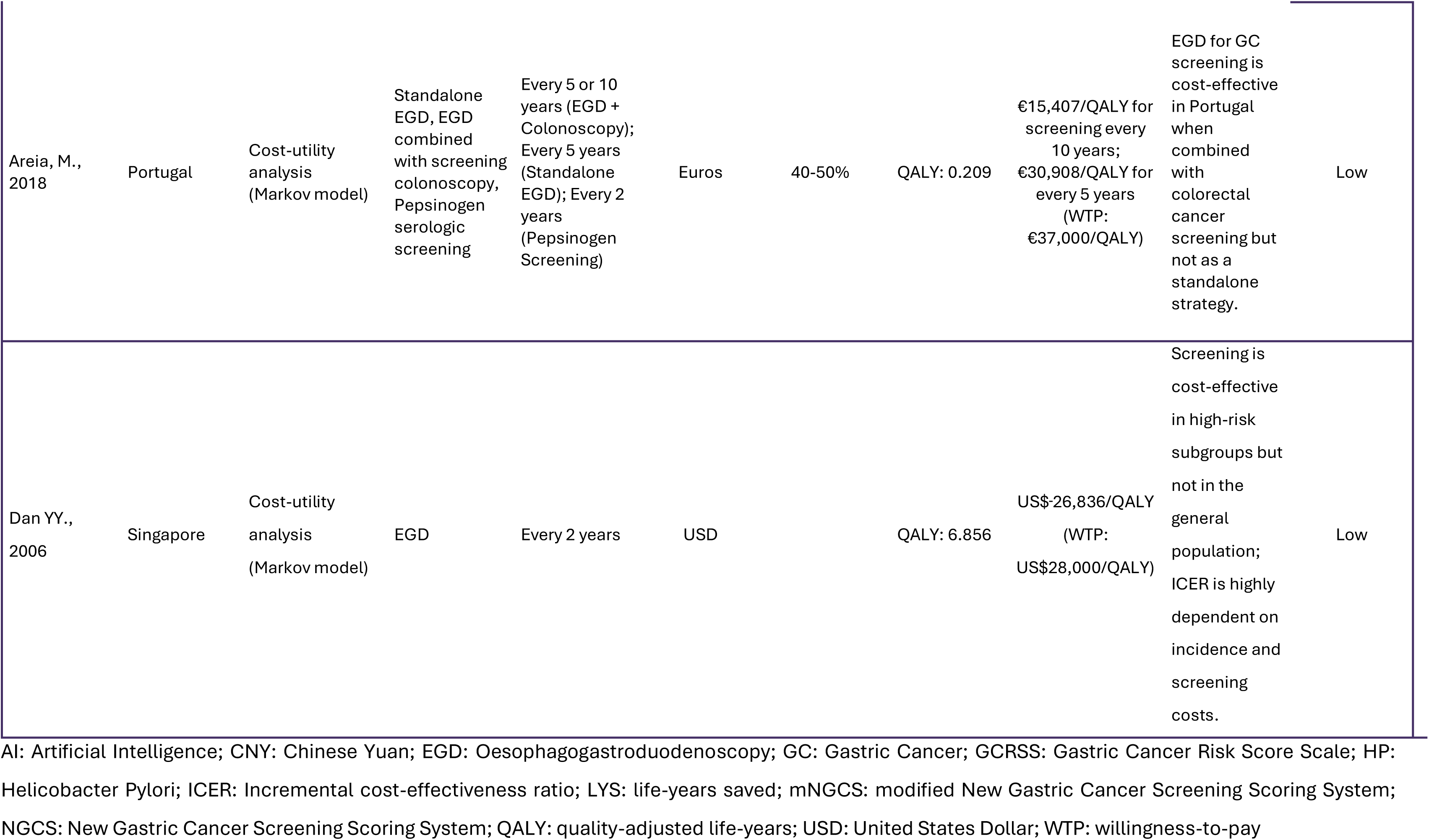
Characteristics of included cost-effectiveness studies and a description of the main outcomes.

| First Author, Year | Country | Type of Study | Type(s) of Screening | Periodicity | Currency | Relative risk reduction of RR of GC mortality | Additional LYS and/or QALY for best screening strategy | ICERs for best screening strategy | Conclusion | Risk of bias |
| --- | --- | --- | --- | --- | --- | --- | --- | --- | --- | --- |
| Libânio, D., 2024 | Portugal | Cost-Effectiveness (Markov model) | Single EGD at 50 years, EGD + Colonoscopy (once or twice per decade), EGD alone (once or twice per decade) with or without AI | Once, every 5 or 10 years | Euros | 60-70% |  | €33,297/QALY for screening every 10 years; €42,214/QALY for every 5 years (WTP: €47,460/QALY) | EGD + Colonoscopy twice per decade is cost-effective in Portugal | Low |
| Qin, S., 2024 | China | Cost-Effectiveness (Microsimulation model) | Endoscopic screening every 2, 3, 5, or 8 years; NGCS; GCRSS | Every 2, 3, 5, or 8 years; NGCS and GCRSS strategies | USD | | NGCS Strategy (40–60 years, no comorbidity): LYS: ~1.13 / QALY: ~1.14 | US\$17,618/QALY (WTP: US\$18,575/QALY) | Personalized screening based on comorbidity is cost-effective; NGCS provides the best balance of benefits and costs | Low |
| Qin, S., 2022 | China | Cost-Effectiveness (Microsimulation model) | GCRSS screening strategies starting at 40, 45, 50, 55, 60, 65, and 70 years | Once, every 2 or 3 years | USD | 70.6% | 40-GCRSS: LYS: 0.795 / QALY: 0.068 | US\$12,601/QALYs (WTP: US\$37,655/QALY) | GCRSS screening from age 40 is cost-effective and reduces GC burden in China | Moderate- |
| Qin, S., 2022 | China | Cost-Effectiveness (Microsimulation and Markov model) | mNGCS, endoscopic screening (once, every 2 years, every 3 years) | Once, every 2 or 3 years | USD | 90.2% | 40-NGCS: LYS: 0.120 /QALY: 0.090 | US\$15,668/QALYs (WTP: US\$17,922/QALY) | NGCS-based screening from 40 years is cost-effective in China, reducing GC burden significantly | Low |
| Wang Z., 2022 | China | Cost-Effectiveness (Markov model) | HP eradication; Endoscopic screening (one-time, annual, biennial, triennial); Risk-stratified screening | Once, every 2 or 3 years | CNY | 44.5% | QALY: 11.99 | CNY 17,725 (WTP:70,000 CNY/QALY) | HP eradication should be prioritized in national health plans; targeted EGD is preferable over general population screening | Low |
| Xia, R., 2021 | China | Cost-Effectiveness (Markov model) | EGD (one-time, every 2, 3, 5, or 10 years) | Once, every 2, 3, 5, or 10 years | USD | | QALY: 1.087 to 10.362 | US\$1,087 to US\$4,511/QALY (WTP: US\$10,276/QALY) | EGD screening for GC and esophageal cancer every 2 years is cost-effective in high-risk areas | Low |
| Areia, M.,<br>2018 | Portugal | Cost-utility<br>analysis<br>(Markov model) | Standalone<br>EGD, EGD<br>combined<br>with screening<br>colonoscopy,<br>Pepsinogen<br>serologic<br>screening | Every 5 or 10<br>years (EGD +<br>Colonoscopy);<br>Every 5 years<br>(Standalone<br>EGD); Every 2<br>years<br>(Pepsinogen<br>Screening) | Euros | 40-50% | QALY: 0.209 | €15,407/QALY for<br>screening every<br>10 years;<br>€30,908/QALY for<br>every 5 years<br>(WTP:<br>€37,000/QALY) | EGD for GC<br>screening is<br>cost-effective<br>in Portugal<br>when<br>combined<br>with<br>colorectal<br>cancer<br>screening but<br>not as a<br>standalone<br>strategy. | Low |
| Dan YY.,<br>2006 | Singapore | Cost-utility<br>analysis<br>(Markov model) | EGD | Every 2 years | USD | | QALY: 6.856 | US\$26,836/QALY<br>(WTP:<br>US\$28,000/QALY) | Screening is<br>cost-effective<br>in high-risk<br>subgroups but<br>not in the<br>general<br>population;<br>ICER is highly<br>dependent on<br>incidence and<br>screening<br>costs. | Low |

AI: Artificial Intelligence; CNY: Chinese Yuan; EGD: Oesophagogastrroduodenoscopy; GC: Gastric Cancer; GCRSS: Gastric Cancer Risk Score Scale; HP: Helicobacter Pylori; ICER: Incremental cost-effectiveness ratio; LYS: life-years saved; mNGCS: modified New Gastric Cancer Screening Scoring System; NGCS: New Gastric Cancer Screening Scoring System; QALY: quality-adjusted life-years; USD: United States Dollar; WTP: willingness-to-pay

Two studies employed microsimulation models^56,57^, while the remaining six adopted variations of Markov model, including cost-utility analysis^13,58^, economic evaluation ^2,73^, and decision analysis.^61^ Notably, one study integrated both Markov and microsimulation approaches^62^, leveraging the strengths of each to model disease progression and cost-effectiveness more comprehensively.

Regarding geographical distribution, five of the studies were conducted in China^56,57,60–62^, indicating a strong research focus on economic evaluations of screening strategies within the Chinese healthcare system. Portugal contributed two studies^13,59^, reflecting a growing interest in cost-effectiveness modelling in European healthcare settings, while Singapore accounted for one study^58^, further emphasizing the international scope of these modelling approaches.

The screening strategies assessed were diverse, ranging from one-time endoscopic examinations to biennial, triennial, and risk-based screening. Some studies also evaluated the cost-effectiveness of HP eradication, either as a standalone strategy or in combination with endoscopic screening. The frequency of screening varied significantly, depending on the target population’s risk level and country-specific economic considerations.

One of the primary outcomes analysed was the reduction in the relative risk of GC mortality. Reported reductions ranged from 40% to 90%, depending on the strategy and population characteristics. The greatest impact was observed in a Chinese study ^62^ that used a Markov model combined with microsimulation and indicated that the implementation of the New Gastric Cancer Screening <u>s</u>ystem starting at age 40 could reduce mortality by up to 90.2% in that population. The same study highlights a reduction in GC incidence up to 86.4% with the same screening strategy.

The impact of screening on survival was assessed in life-years saved and/or quality-adjusted life-years (QALY), and the cost-effectiveness analysis was assessed by incremental cost-effectiveness ratio (ICER) concerning the willingness-to-pay (WTP) of each country.

Despite reporting cost-effectiveness outcomes, most studies did not provide confidence intervals or standard errors for life-years saved, QALY, or ICER.

### Quality-Adjusted Life Years (Qaly)

The highest reported QALY gain was observed in Wang et al.^61^, where a combined strategy of HP eradication and EGD screening resulted in an estimated gain of 11.990 QALYs, representing the most beneficial intervention in terms of health outcomes. The analysis by Xia et al.^60^ and Dan et al^58^ showed that the strategy with the greatest QALY gain (10.36 and 6.86 years per person, respectively) is EGD screening every 2 years between the ages of 40 and 44, reinforcing the effectiveness of frequent endoscopic surveillance in high-risk settings. Qin et al.^56^ analysed personalised risk-based screening, incorporating comorbidities and age, reporting an estimated gain of 1.140 QALYs, demonstrating that individualised screening schedules can yield substantial health benefits. Libânio et al.^59^, which assessed EGD and colonoscopy without and with artificial intelligence (AI) every 5 and 10 years reports that combined screening twice per decade yields the highest effectiveness with a QALY of 1.12 years per person. Areia et al.^13^ evaluated EGD screening every 5 and 10 years in combination with colorectal cancer screening and reported a QALY gain of 0.209, reflecting a benefit compared with no screening.

Another 2 studies of Qin et al.^57,62^ examined different risk-stratified screening strategies, reporting that New Gastric Cancer Screening system, screening from age 40 yielded 0.090 additional QALYs, while Gastric Cancer Risk Score Scale based screening resulted in 0.068 QALYs, highlighting the potential advantages of risk-stratified screening over uniform population-based approaches.

### Incremental Cost-Effectiveness Ratio (Icer)

ICER values varied considerably depending on the screening method, target population, and healthcare setting. Currency was standardised to United States dollars (US$) at the February 2025 exchange rate (1€=1.04US$ and 1CN=0.138US$) to facilitate comparisons between studies. The lowest ICER was observed in Wang et al.^61^, where HP eradication resulted in an ICER of US$2,444/QALY, well below the WTP threshold of US$9,654/QALY, confirming its cost-effectiveness. Xia et al.^60^ found that biennial endoscopic screening in high-risk areas had an ICER ranging from US$1,087 to US$4,511/QALY, the most cost-efficient among endoscopic screening approaches, given the WTP threshold of US$10,276/QALY in the study’s setting. Qin et al.^57,62^ reported that Gastric Cancer Risk Score Scale screening from age 40 had an ICER of US$12,601/QALY, remaining well within the US$37,655/QALY WTP limit, while the New Gastric Cancer Screening strategy was associated with an ICER of US$15,668/QALY, falling below the US$17,922/QALY threshold. Another study from the same authors^56^ found that personalised risk-based screening had an ICER of US$17,618/QALY, aligning with the US$18,575/QALY WTP threshold, reinforcing its cost-effectiveness compared to universal screening strategies. Areia et al.^13^ reported two different ICER values based on screening frequency: endoscopic screening every 10 years combined with colorectal cancer screening had an ICER of US$16,023/QALY, whereas screening every 5 years had an ICER of US$32,144/QALY, both of which were within Portugal’s US$38,480/QALY WTP threshold. Finally, Libânio et al.^59^, which assessed EGD combined with colonoscopy every 5 or 10 years, reported an ICER of US$34,629/QALY and US$43,903/QALY, respectively, both remaining within the US$49,358/QALY WTP threshold. The same author reported that combined EGD and colonoscopy with AI twice per decade strategy is cost-effective as long as AI accuracy is at least 93% and the AI cost per endoscopy is ≤ US$20.8.

## DISCUSSION

This systematic review and meta-analysis provide updated evidence on the effectiveness and cost-effectiveness of endoscopic screening for GC in intermediate-risk countries. The findings indicate that endoscopic screening significantly increases the detection of precancerous conditions and early-stage GC, with a pooled early detection rate of 73.6%, aligning with results from high-risk countries such as Japan and South Korea, where national screening programmes have demonstrated a similar stage shift.^11,63^ Similarly, Khanderia et al.^64^ demonstrated that screening nearly quadruples the likelihood of early-stage detection (odds ratio 3.90, 95% CI 3.01–5.06), with corresponding improvements in five-year survival rates.

Substantial heterogeneity was observed across studies, likely due to differences in design, screening methodology, and population characteristics. A major contributor to this variability is the predominance of studies from China. Although classified as intermediate-risk, some Chinese regions, particularly rural areas, have incidence rates comparable to high-risk settings. Two large-scale screening programmes, targeting rural and urban populations, accounted for a significant proportion of the included studies. This geographic disparity may have influenced detection rates and effectiveness estimates. Furthermore, pre-screening strategies such as HP testing and risk stratification were employed in several studies, potentially improving screening efficiency and impacting overall outcomes.

Screening’s impact extends beyond early detection. Included studies reported mortality reduction from 40% to 90%, depending on the screening strategy and population. The greatest benefit was seen in risk-stratified approaches, such as the New Gastric Cancer Screening System, which reduced mortality by up to 90.2% when initiated at age 40. These findings are aligned with previous meta-analyses, including Hibino et al.^65^, who demonstrated a significant mortality reduction (relative risk of 0.52, 95% CI 0.39–0.79).

The effectiveness also depends on interval and adherence. Included studies evaluated a wide range of screening frequencies, from a single endoscopic examination to biennial, triennial, and risk-based approaches. Shorter intervals were generally associated with higher early detection rates. However, adherence varied widely across studies (12.7% to 94.2%), suggesting that participation rates may significantly impact real-world effectiveness. In addition, HP infection plays a crucial role in GC risk, with prevalence varying significantly across geographic regions. In this review, HP infection rates ranged from 13.1% to 89.2% in this review (pooled effect size of 49.4%). Hooi et al.^66^ reported similar global variations, with rates exceeding 70% in some Asian regions but falling below 30% in Oceania. Some studies evaluated HP eradication as a complementary or alternative strategy to endoscopic screening. Morais et al.^67^ found that despite declining GC mortality in Portugal, HP prevalence remains elevated among older age groups, underscoring the potential value of combined eradication and screening strategies.

Cost-effectiveness is a critical determinant in the feasibility of implementing GC screening programmes, particularly in intermediate-risk settings where resource allocation must be balanced with expected health benefits. The eight studies included in this review employed microsimulation and Markov modelling techniques, widely recognised for their applicability in economic evaluations. Markov models predominated, reflecting their common use in modelling disease progression and healthcare costs. Microsimulation was utilised in two studies, allowing for more individualised analyses, while one study integrated both methodologies. A primary outcome of these studies was the reduction in GC mortality risk, which ranged from 40% to 90%. The highest impact was observed in a Chinese study that used a Markov-microsimulation model, demonstrating that the New Gastric Cancer Screening System initiated at the age of 40 could reduce mortality by 90.2% and incidence by up to 86.4%.^62^ These findings align with cost-effectiveness evaluations from Japan and South Korea, where national screening programmes have been shown to reduce GC mortality.^68,69^ In Japan, the implementation of systematic screening programs has led to a significant increase in the five-year survival rate, ranging between 50% and 70%, attributed to early detection enabled by these programmes.^63^ In South Korea, the National Cancer Screening Programme demonstrated that individuals undergoing regular endoscopic screenings had a 47% reduction in GC mortality risk compared to those who had never been screened (odds ratio 0.53; 95% CI 0.51–0.56).^70^ Additionally, the five-year survival rate in South Korea exceeds 65%, reinforcing the effectiveness of screening programmes in reducing mortality.^70^

The economic benefits of screening were assessed through life-years saved and QALY. The highest QALY gains were reported by Wang et al.^61^, where a combined strategy of HP eradication and endoscopic screening resulted in a gain of 11.99 QALYs. Xia et al.^60^ and Dan et al.^58^ found that biennial endoscopic screening in high-risk populations provided substantial QALY gains (10.36 and 6.86 years per person, respectively), reinforcing the cost-effectiveness of frequent surveillance. Incremental ICER estimates varied depending on the screening approach and healthcare setting. Wang et al.^61^ found that HP eradication had the lowest ICER (US$2,444/QALY), well below the WTP threshold. Xia et al.^60^ demonstrated that biennial endoscopic screening in high-risk areas had an ICER ranging from US$1,087 to US$4,511/QALY, confirming its economic viability. Several studies by Qin et al.^56,57,62^ reported that risk-stratified screening models remained within acceptable WTP thresholds. European cost-effectiveness analyses further support targeted screening approaches. Areia et al.^13^ reported that endoscopic screening every 10 years combined with colorectal cancer screening had an ICER of US$16,023/QALY, whereas screening every five years had an ICER of US$32,144/QALY, both within Portugal’s WTP threshold. Libânio et al.^59^ demonstrated that EGD combined with colonoscopy remains cost-effective, provided AI accuracy exceeds 93% and AI costs per endoscopy remain below US$20.8. These results highlight the potential of AI-enhanced screening to optimise resource use and outcomes.

Importantly, the results of this systematic review are in line with the recently published Management of epithelial precancerous conditions and early neoplasia of the stomach III guidelines, issued by the European Society of Gastrointestinal Endoscopy in 2025, which state that endoscopic screening may be considered in intermediate-risk countries, provided that cost-effectiveness has been demonstrated.^16^

## LIMITATIONS

Despite the robust findings presented in this review, several limitations should be acknowledged. First, substantial heterogeneity was observed among the included studies, likely due to differences in study design, screening methodologies, and population characteristics. The predominance of studies from China, where GC incidence varies significantly between urban and rural areas, may have influenced the overall estimates of screening effectiveness. Additionally, variations in pre-screening strategies, such as the use of HP testing and risk stratification models, may contribute to inconsistencies in reported outcomes. Another limitation is the variation in adherence rates, which ranged from 12.7% to 94.2% across studies. Since participation rates can significantly affect the real-world effectiveness of screening programmes, this variability may limit the generalisability of findings.

Concerning cost-effectiveness analyses, several limitations should also be considered. Differences in modelling approaches, particularly between Markov models and microsimulations, may introduce variability in cost estimates. Additionally, healthcare costs and economic thresholds vary significantly across countries, making it difficult to generalise findings to different healthcare systems. Many studies assume high adherence rates and optimal implementation conditions, which may not reflect real-world scenarios where participation is inconsistent. Moreover, the long-term economic impact of integrating AI into endoscopic screening remains uncertain, as its cost-effectiveness depends on factors such as diagnostic accuracy and implementation costs. Lastly, while cost-effectiveness models often rely on projections, there is a need for more long-term empirical data to validate these estimates and ensure their applicability across diverse populations.

Finally, it is important to acknowledge that other intermediate-risk countries may be conducting research on endoscopic screening for gastroesophageal cancer that has not yet been published or included in the current literature. This potential publication lag may result in an incomplete representation of ongoing screening initiatives, emphasising the need for continuous updates and broader inclusion of emerging evidence.

## IMPLICATIONS FOR FUTURE RESEARCH

Future studies should focus on conducting well-designed randomised trials to establish more robust evidence on the benefits and feasibility of endoscopic screening in intermediate-risk settings. Standardising screening protocols, including risk stratification and adherence strategies, will be essential to improving comparability between studies. Additionally, economic evaluations tailored to different healthcare environments are needed to determine the feasibility of large-scale implementation.

Research on innovative screening technologies, such as AI-assisted endoscopy and non-invasive biomarkers, could further optimise cost-effectiveness by reducing costs and increasing diagnostic efficiency. Long-term impact assessments of screening on GC mortality and healthcare expenditures will also be crucial in guiding public health policies.

## CONCLUSION

This systematic review provides strong evidence supporting the effectiveness and cost-effectiveness of endoscopic screening for GC in intermediate-risk countries. Screening programmes have been shown to significantly improve early detection rates and reduce GC mortality, particularly in settings where risk-stratified approaches are implemented. The findings align with experiences from high-risk countries such as Japan and South Korea, reinforcing the potential benefits of adopting similar strategies in intermediate-risk settings.

Despite these positive outcomes, important challenges remain, including study heterogeneity, variability in adherence rates, and uncertainties in cost-effectiveness modelling. Future research should focus on optimising screening protocols, improving participation rates, and addressing economic feasibility in diverse healthcare systems. Additionally, continuous monitoring and updates to the literature will be essential to incorporating new evidence from ongoing screening initiatives worldwide.

## LIST OF ABBREVIATIONS

AI: Artificial Intelligence
CI: Confidence Interval
EGC: Early Gastric Cancer
EGD: Esophagogastroduodenoscopy
GC: Gastric Cancer
HP: Helicobacter pylori
ICER: Incremental Cost-Effectiveness Ratio
QALY: Quality-Adjusted Life-Year
WTP: Willingness-To-Pay

## Data Availability

All data generated or analysed during this study are included in this published article (and its supplementary information files).

## ACKNOWLEDGMENTS

This manuscript is part of author Mourato B.’s PhD project in Health and Well-being Sciences and Technologies and will be integrated into her final thesis.

## AUTHORS’ CONTRIBUTIONS

BM was responsible for conceptualization, designing, investigation, formal analysis and writing the manuscript.

NP provided assistance with protocol design, investigation, and writing.

ABP was responsible for investigation, and reviewing the writing of the manuscript. RC was responsible for designing the search expression in the various databases. IF and MA participated in protocol design, systematic review process supervision, and manuscript review.

RD was responsible for the revision of the manuscript.

## DECLARATIONS OF AI USE

During the preparation of this work the authors used ChatGPT (OpenAI) in order to assist with language refinement. After using this tool, the authors reviewed and edited the content as needed and take full responsibility for the content of the publication.

## SUPLEMENTARY MATERIAL

Supplementary File 1 - Search strategy.docx Supplementary File 2 – variables form. docx Supplementary File 3 - All Studies Rayyan.xlsx Supplementary File 4 – Excluded Studies.xlsx Supplementary File 5 - Included Studies Dataset.xlsx

Supplementary File 6 - Included studies outcomes dataset.xlsx Supplementary File 7 - Helicobacter Pylori Meta-analysis.pdf Supplementary File 8 - Precancerous lesions Meta-analysis.pdf Supplementary File 9 - Positive lesions Meta-analysis.pdf Supplementary File 10 - EGC Meta-analysis.pdf

## REFERENCES

1. International Agency for Research on Cancer. Global Cancer Observatory 2022 - Stomach. Published online 2022. https://gco.iarc.fr/today/en/dataviz/maps-heatmap?mode=population&cancers=7

2. Cristina Díaz del Arco, Luis Ortega Medina, Lourdes Estrada Muñoz, Soledad García Gómez de las Heras and Maria Jesús Fernández Aceñero. Is there still a place for conventional histopathology in the age of molecular medicine? Laurén classification, inflammatory infiltration and other current topics in gastric cancer diagnosis and prognosis. Histol Histopathol. 2021;36(6):587-613. doi:10.14670/HH-18-309

3. Arnold M, Park JY, Camargo MC, Lunet N, Forman D, Soerjomataram I. Is gastric cancer becoming a rare disease? A global assessment of predicted incidence trends to 2035. Gut. 2020;69(5):823- 829. doi:10.1136/gutjnl-2019-320234

4. Bernardes A, ed. Carcinoma gástrico. 1. ͣ edição. Lidel; 2021.

5. Januszewicz W, Turkot MH, Malfertheiner P, Regula J. A Global Perspective on Gastric Cancer Screening: Which Concepts Are Feasible, and When? Cancers. 2023;15(3):664. doi:10.3390/cancers15030664

6. Allemani C, Matsuda T, Di Carlo V, et al. Global surveillance of trends in cancer survival 2000–14 (CONCORD-3): analysis of individual records for 37 513 025 patients diagnosed with one of 18 cancers from 322 population-based registries in 71 countries. The Lancet. 2018;391(10125):1023–1075. doi:10.1016/S0140-6736(17)33326-3

7. Suzuki H, Oda I, Abe S, et al. High rate of 5-year survival among patients with early gastric cancer undergoing curative endoscopic submucosal dissection. Gastric Cancer. 2016;19(1):198–205. doi:10.1007/s10120-015-0469-0

8. Necula L, Matei L, Dragu D, et al. Recent advances in gastric cancer early diagnosis. WJG. 2019;25(17):2029–2044. doi:10.3748/wjg.v25.i17.2029

9. Vradelis S, Maynard N, Warren BF, Keshav S, Travis SPL. Quality control in upper gastrointestinal endoscopy: detection rates of gastric cancer in Oxford 2005–2008. Postgraduate Medical Journal. 2011;87(1027):335-339. doi:10.1136/pgmj.2010.101832

10. Hamashima C. Cancer screening guidelines and policy making: 15 years of experience in cancer screening guideline development in Japan. Japanese Journal of Clinical Oncology. 2018;48(3):278–286. doi:10.1093/jjco/hyx190

11. Suh Y, Lee J, Woo H, et al. National cancer screening program for gastric cancer in Korea: Nationwide treatment benefit and cost. Cancer. 2020;126(9):1929–1939. doi:10.1002/cncr.32753

12. Lansdorp-Vogelaar I. Cost-effectiveness of prevention and early detection of gastric cancer in Western countries. Best Practice. 2021;Volumes 50–51.

13. Areia M, Spaander MC, Kuipers EJ, Dinis-Ribeiro M. Endoscopic screening for gastric cancer: A cost-utility analysis for countries with an intermediate gastric cancer risk. UEG Journal. 2018;6(2):192–202. doi:10.1177/2050640617722902

14. Burra P, Bretthauer M, Buti Ferret M, et al. Digestive cancer screening across Europe. UEG Journal. 2022;10(4):435–437. doi:10.1002/ueg2.12230

15. Săftoiu A, Hassan C, Areia M, et al. Role of gastrointestinal endoscopy in the screening of digestive tract cancers in Europe: European Society of Gastrointestinal Endoscopy (ESGE) Position Statement. Endoscopy. 2020;52(04):293–304. doi:10.1055/a-1104-5245

16. Dinis-Ribeiro M, Libânio D, Uchima H, et al. Management of epithelial precancerous conditions and early neoplasia of the stomach (MAPS III): European Society of Gastrointestinal Endoscopy (ESGE), European Helicobacter and Microbiota Study Group (EHMSG) and European Society of Pathology (ESP) Guideline update 2025. Endoscopy. Published online March 20, 2025:a-2529- 5025. doi:10.1055/a-2529-5025

17. Mourato MB, Pratas N, Branco Pereira A, et al. Effectiveness of Gastric Cancer Endoscopic Screening in Intermediate-Risk Countries: Protocol for a Systematic Review and Meta-Analysis. JMIR Res Protoc. 2025;14:e56791. doi:10.2196/56791

18. McGowan J, Sampson M, Salzwedel DM, Cogo E, Foerster V, Lefebvre C. PRESS Peer Review of Electronic Search Strategies: 2015 Guideline Statement. Journal of Clinical Epidemiology. 2016;75:40–46. doi:10.1016/j.jclinepi.2016.01.021

19. Wells, G, Shea, B, O’Connell, D, et al. The Newcastle-Ottawa Scale (NOS) for Assessing the Quality of Nonrandomised Studies in Meta-Analyses. Published online 2011.

20. National Heart, Lung and Blood Institute. Study Quality Assessment Tools. https://www.nhlbi.nih.gov/health-topics/study-quality-assessment-tools

21. Evers S, Goossens M, De Vet H, Van Tulder M, Ament A. Criteria list for assessment of methodological quality of economic evaluations: Consensus on Health Economic Criteria. Int J Technol Assess Health Care. 2005;21(2):240–245. doi:10.1017/S0266462305050324

22. Schlemper RJ. The Vienna classification of gastrointestinal epithelial neoplasia. Gut. 2000;47(2):251–255. doi:10.1136/gut.47.2.251

23. Borenstein, M., Hedges, L.V., Higgins, J.P.T. and Rothstein, H.R. *Introduction to Meta-Analysis*. Wiley.; 2021. Accessed March 4, 2025. 10.1002/9780470743386.fmatter

24. Edwards AWF. Chapter 67 - R.A. Fischer, statistical methods for research workers, first edition (1925). In: Grattan-Guinness I, Cooke R, Corry L, Crépel P, Guicciardini N, eds. Landmark Writings in Western Mathematics 1640-1940. Elsevier Science; 2005:856-870. doi:10.1016/B978-044450871-3/50148-0

25. Higgins JPT. Measuring inconsistency in meta-analyses. BMJ. 2003;327(7414):557-560. doi:10.1136/bmj.327.7414.557

26. Veroniki AA, Jackson D, Viechtbauer W, et al. Methods to estimate the between-study variance and its uncertainty in meta-analysis. Research Synthesis Methods. 2016;7(1):55–79. doi:10.1002/jrsm.1164

27. Landis JR, Koch GG. The Measurement of Observer Agreement for Categorical Data. Biometrics. 1977;33(1):159. doi:10.2307/2529310

28. Cohen J. A Coefficient of Agreement for Nominal Scales. Educational and Psychological Measurement. 1960;20(1):37–46. doi:10.1177/001316446002000104

29. Zeng H, Sun K, Cao M, et al. Initial results from a multi-center population-based cluster randomized trial of esophageal and gastric cancer screening in China. BMC Gastroenterol. 2020;20(1):398. doi:10.1186/s12876-020-01517-3

30. Zheng X, Mao X, Xu K, et al. Massive Endoscopic Screening for Esophageal and Gastric Cancers in a High-Risk Area of China. Green J, ed. PLoS ONE. 2015;10(12):e0145097. doi:10.1371/journal.pone.0145097

31. Ge X, Zhang X, Ma Y, Chen S, Chen Z, Li M. Diagnostic value of macrophage inhibitory cytokine 1 as a novel prognostic biomarkers for early gastric cancer screening. Clinical Laboratory Analysis. 2021;35(1):e23568. doi:10.1002/jcla.23568

32. Chen Q, Yu L, Hao C, et al. Effectiveness of endoscopic gastric cancer screening in a rural area of Linzhou, China: results from a case–control study. Cancer Medicine. 2016;5(9):2615–2622. doi:10.1002/cam4.812

33. Chen R, Liu Y, Song G, et al. Effectiveness of one-time endoscopic screening programme in prevention of upper gastrointestinal cancer in China: a multicentre population-based cohort study. Gut. Published online April 2, 2020:gutjnl-2019-320200. doi:10.1136/gutjnl-2019-320200

34. Riecken B, Pfeiffer R, Ma JL, et al. No Impact of Repeated Endoscopic Screens on Gastric Cancer Mortality in a Prospectively Followed Chinese Population at High Risk. Preventive Medicine. 2002;34(1):22–28. doi:10.1006/pmed.2001.0925

35. Xiao H, Yan S, Li J, et al. Development and external validation of a nomogram to predict the risk of Upper gastrointestinal precancerous lesions in a non-high-incidence area. Cancer Medicine. 2020;9(22):8722–8732. doi:10.1002/cam4.3462

36. Tong Y, Wang H, Zhao Y, et al. Diagnostic Value of Serum Pepsinogen Levels for Screening Gastric Cancer and Atrophic Gastritis in Asymptomatic Individuals: A Cross-Sectional Study. Front Oncol. 2021;11:652574. doi:10.3389/fonc.2021.652574

37. Emura F, Mejía J, Mejía M, et al. Effectiveness of systematic chromoendoscopy for diagnosis of early cancer and gastric premalignant lesions. Results of two consecutive screening campaigns in Colombia (2006-2007).

38. Zhou X, Zhu H, Zhu C, et al. Helicobacter pylori Infection and Serum Pepsinogen Level With the Risk of Gastric Precancerous Conditions: A Cross-sectional Study of High-risk Gastric Cancer Population in China. Journal of Clinical Gastroenterology. 2021;55(9):778–784. doi:10.1097/MCG.0000000000001444

39. Li WQ, Qin XX, Li ZX, et al. Beneficial effects of endoscopic screening on gastric cancer and optimal screening interval: a population-based study. Endoscopy. 2022;54(09):848–858. doi:10.1055/a-1728-5673

40. Li L, Ni J, Sun S, Zha X, Li R, He C. Clinical applicability of a new scoring system for population-based screening and risk factors of gastric cancer in the Wannan region. BMC Gastroenterol. 2022;22(1):306. doi:10.1186/s12876-022-02384-w

41. Ji L, Liu Z, Zhou B, et al. Community-Based Pilot Study of a Screening Program for Gastric Cancer in a Chinese Population. Cancer Prevention Research. 2020;13(1):73–82. doi:10.1158/1940-6207.CAPR-19-0372

42. Yu Z, Zuo T, Yu H, et al. Outcomes of upper gastrointestinal cancer screening in high-risk individuals: a population-based prospective study in Northeast China. BMJ Open. 2022;12(2):e046134. doi:10.1136/bmjopen-2020-046134

43. Huang Y, Li H, Long X, Liang X, Lu H. Lessons learned from upper gastrointestinal endoscopy in asymptomatic Chinese. Helicobacter. 2021;26(3):e12803. doi:10.1111/hel.12803

44. Liu Y, Gu K. Association between anesthesia assistance and precancerous lesions and early cancer detection during diagnostic esophagogastroduodenoscopy: a propensity score-matched retrospective study. Front Med. 2024;11:1389809. doi:10.3389/fmed.2024.1389809

45. Meng T, Wu Y, Chen X, Lu J, Fan J, Bai J. Results of endoscopy in 35,525 patients with precancerous diseases of the gastrointestinal tract.

46. Mansour-Ghanaei F, Sokhanvar H, Joukar F, et al. Endoscopic Findings in a Mass Screening Program for Gastric Cancer in a High Risk Region - Guilan Province of Iran. Asian Pacific Journal of Cancer Prevention. 2012;13(4):1407–1412. doi:10.7314/APJCP.2012.13.4.1407

47. Malekzadeh R. Prevalence of gastric precancerous lesions in Ardabil, a high incidence province for gastric adenocarcinoma in the northwest of Iran. Journal of Clinical Pathology. 2004;57(1):37–42. doi:10.1136/jcp.57.1.37

48. Dorji T, Wangmo S, Dargay S, et al. Population-level cancer screening and cancer care in Bhutan, 2020–2023: a review. The Lancet Regional Health - Southeast Asia. 2024;24:100370. doi:10.1016/j.lansea.2024.100370

49. Machaca Quea N, Emura F, Barreda Bolaños F, Salvador Arias Y, Arévalo Suárez F, Piscoya Rivera A. Effectiveness of systematic alphanumeric coded endoscopy for diagnosis of gastric intraepithelial neoplasia in a low socioeconomic population. Endosc Int Open. 2016;04(10):E1083–E1089. doi:10.1055/s-0042-115408

50. Morais R, Moreira J, Gaspar R, et al. Higher frequency of gastric neoplasia in advanced chronic liver disease patients: Impact of screening endoscopy in an intermediate-high risk country. Digestive and Liver Disease. Published online May 2024:S1590865824007345. doi:10.1016/j.dld.2024.04.035

51. Lau JWL, Khoo MJW, Leong XH, et al. Opportunistic upper endoscopy during colonoscopy as a screening strategy for countries with intermediate gastric cancer risk. J of Gastro and Hepatol. 2021;36(4):1081–1087. doi:10.1111/jgh.15290

52. Zheng X, Mao X, Xu K, et al. Massive Endoscopic Screening for Esophageal and Gastric Cancers in a High-Risk Area of China. Green J, ed. PLoS ONE. 2015;10(12):e0145097. doi:10.1371/journal.pone.0145097

53. Xiao HF, Yan SP, Chen Y fang, et al. Community-Based Upper Gastrointestinal Cancer Screening in a Randomized Controlled Trial: Baseline Results in a Non-high-incidence Area. Cancer Prevention Research. 2020;13(3):317–328. doi:10.1158/1940-6207.CAPR-19-0422

54. Gong Y, Kang J, Wu R, Ge F, Zheng Y song, Zeng Q. Gastroscopic results for the asymptomatic, average-risk population in Northern China: a cross-sectional study of 60,519 adults. Scandinavian Journal of Gastroenterology. Published online February 8, 2022:1–9. doi:10.1080/00365521.2022.2035810

55. Meng T, Wu Y, Chen X, Lu J, Fan J, Bai J. Results of endoscopy in 35,525 patients with precancerous diseases of the gastrointestinal tract.

56. Qin S, Wang X, Li S, Wu M, Wan X. Personalizing age of gastric cancer screening based on comorbidity in China: Model estimates of benefits, affordability and cost-effectiveness optimization. Preventive Medicine. 2024;179:107851. doi:10.1016/j.ypmed.2024.107851

57. Qin S, Wang X, Li S, et al. Benefit-to-harm ratio and cost-effectiveness of government-recommended gastric cancer screening in China: A modeling study. Front Public Health. 2022;10:955120. doi:10.3389/fpubh.2022.955120

58. Dan YY, So JBY, Yeoh KG. Endoscopic Screening for Gastric Cancer. Clinical Gastroenterology and Hepatology. 2006;4(6):709–716. doi:10.1016/j.cgh.2006.03.025

59. Libanio D, Antonelli G, Marijnissen F, et al. Combined gastric and colorectal cancer endoscopic screening may be cost-effective in Europe with the implementation of artificial intelligence: an economic evaluation. European Journal of Gastroenterology & Hepatology. 2024;36(2):155–161. doi:10.1097/MEG.0000000000002680

60. Xia R, Zeng H, Liu W, et al. Estimated Cost-effectiveness of Endoscopic Screening for Upper Gastrointestinal Tract Cancer in High-Risk Areas in China. JAMA Netw Open. 2021;4(8):e2121403. doi:10.1001/jamanetworkopen.2021.21403

61. Wang Z, Han W, Xue F, et al. Nationwide gastric cancer prevention in China, 2021–2035: a decision analysis on effect, affordability and cost-effectiveness optimisation. Gut. 2022;71(12):2391-2400. doi:10.1136/gutjnl-2021-325948

62. Qin S, Wang X, Li S, et al. Clinical Benefit and Cost Effectiveness of Risk-Stratified Gastric Cancer Screening Strategies in China: A Modeling Study. PharmacoEconomics. 2022;40(7):725–737. doi:10.1007/s40273-022-01160-8

63. Narii N, Sobue T, Zha L, et al. Effectiveness of endoscopic screening for gastric cancer: The Japan Public Health Center-based Prospective Study. Cancer Science. 2022;113(11):3922–3931. doi:10.1111/cas.15545

64. Khanderia E, Markar SR, Acharya A, Kim Y, Kim YW, Hanna GB. The Influence of Gastric Cancer Screening on the Stage at Diagnosis and Survival: A Meta-Analysis of Comparative Studies in the Far East. Journal of Clinical Gastroenterology. 2016;50(3):190–197. doi:10.1097/MCG.0000000000000466

65. Hibino M, Hamashima C, Iwata M, Terasawa T. Radiographic and endoscopic screening to reduce gastric cancer mortality: a systematic review and meta-analysis. The Lancet Regional Health - Western Pacific. 2023;35:100741. doi:10.1016/j.lanwpc.2023.100741

66. Hooi JKY, Lai WY, Ng WK, et al. Global Prevalence of Helicobacter pylori Infection: Systematic Review and Meta-Analysis. Gastroenterology. 2017;153(2):420–429. doi:10.1053/j.gastro.2017.04.022

67. Morais S, Ferro A, Bastos A, Castro C, Lunet N, Peleteiro B. Trends in gastric cancer mortality and in the prevalence of Helicobacter pylori infection in Portugal. European Journal of Cancer Prevention. 2016;25(4):275–281. doi:10.1097/CEJ.0000000000000183

68. Hamashima C, Systematic Review Group and Guideline Development Group for Gastric Cancer Screening Guidelines. Update version of the Japanese Guidelines for Gastric Cancer Screening. Japanese Journal of Clinical Oncology. 2018;48(7):673–683. doi:10.1093/jjco/hyy077

69. Kim TH, Kim IH, Kang SJ, et al. Korean Practice Guidelines for Gastric Cancer 2022: An Evidence-based, Multidisciplinary Approach. J Gastric Cancer. 2023;23(1):3. doi:10.5230/jgc.2023.23.e11

70. Hoang T, Woo H, Cho S, Lee J, Kazmi SZ, Shin A. Descriptive Analysis of Gastric Cancer Mortality in Korea, 2000-2020. Cancer Res Treat. 2022;55(2):603-617. doi:10.4143/crt.2022.307

