## Supplementary material 1 for "Effectiveness of Gastric Cancer Endoscopic Screening in Intermediate-Risk Countries – a Systematic Review and Meta-Analysis"

The authors present draft of search strategy to be used in MEDLINE (this search will be adapted to the remaining databases researched):

((Stomach OR Gastric*) AND (Neoplas* OR Cancer OR Tumor* OR Tumour* OR Carcinoma* OR Adenocarcinoma* OR Malignan*) AND (Endoscopy OR Endoscop* OR Gastroscop*) AND (Mass Screening OR Screening*)) AND (tajikistan OR iran OR azerbaijan OR kyrgyzstan OR bhutan OR belarus OR peru OR mali OR chile OR "costa rica" OR china OR kazakhstan OR "russian federation" OR russia OR "vietnam" OR estonia OR colombia OR portugal OR ecuador OR albania OR guadalupe OR turkiye OR lithuania OR lao OR "sao tomé e principe" OR afghanistan OR martinique OR brunei OR zimbabwe OR uzbekistan OR Intermediate Risk OR Moderate Risk OR Intermediate Incidence OR Moderate Incidence OR West* OR Europ*)

Draft of search strategy in SCOPUS:

TITLE-ABS-KEY(Stomach OR Gastric*) AND TITLE-ABS-KEY(Neoplas* OR Cancer OR Tumor* OR Tumour* OR Carcinoma* OR Adenocarcinoma* OR Malignan*) AND TITLE-ABS-KEY(Endoscop* OR Gastroscop*) AND TITLE-ABS-KEY("Mass Screening" OR Screening*) AND TITLE-ABS-KEY("Intermediate Risk" OR "Moderate Risk" OR "Intermediate Incidence" OR "Moderate Incidence" OR West* OR Europ* OR tajikistan OR iran OR azerbaijan OR kyrgyzstan OR bhutan OR belarus OR peru OR mali OR chile OR "costa rica" OR china OR kazakhstan OR "russian federation" OR russia OR vietnam OR estonia OR colombia OR portugal OR ecuador OR albania OR guadalupe OR turkiye OR lithuania OR lao OR "sao tomé e principe" OR afghanistan OR martinique OR brunei OR zimbabwe OR uzbekistan)

Draft of search strategy in EMBASE:

('stomach cancer'/exp OR 'stomach cancer':ti,ab OR 'gastric cancer':ti,ab OR 'gastric malignan*':ti,ab OR 'malignancies of the stomach':ti,ab OR 'malignancy of the stomach':ti,ab OR 'malignant gastric neoplas*':ti,ab OR 'malignant gastric tumor*':ti,ab OR 'malignant neoplasm of the stomach':ti,ab OR 'malignant neoplasms of the stomach':ti,ab OR 'malignant tumor of the stomach':ti,ab OR 'malignant tumors of the stomach':ti,ab OR 'malignant tumour of the stomach':ti,ab OR 'malignant tumours of the stomach':ti,ab OR 'stomach malignan*':ti,ab OR 'stomach tumor'/exp OR 'gastric neoplas*':ti,ab OR 'gastric tumor*':ti,ab OR 'gastric tumour*':ti,ab OR 'neoplasm of the stomach':ti,ab OR 'neoplasms of the stomach':ti,ab OR 'stomach neoplas*':ti,ab OR 'stomach tumor*':ti,ab OR 'stomach tumour*':ti,ab OR 'tumor of the gastric':ti,ab OR 'tumor of the stomach':ti,ab OR 'tumour of the gastric':ti,ab OR 'tumour of the stomach':ti,ab OR 'stomach adenocarcinoma'/exp OR 'gastric adenocarcinoma*':ti,ab OR 'stomach adenocarcinoma*':ti,ab) AND ('endoscopy'/exp OR 'endoscop*':ti,ab OR 'gastroscopy'/exp OR 'gastroscop*':ti,ab) AND ('mass screening'/exp OR 'mass screening':ti,ab OR 'screening':ti,ab) AND ('intermediate risk population'/exp OR 'intermediate risk':ti,ab OR 'moderate risk':ti,ab OR 'intermediate incidence':ti,ab OR 'moderate incidence':ti,ab OR west*:ti,ab OR 'europe'/exp OR 'europ*':ti,ab OR tajikistan:ti,ab OR iran:ti,ab OR azerbaijan:ti,ab OR kyrgyzstan:ti,ab OR bhutan:ti,ab OR belarus:ti,ab OR peru:ti,ab OR mali:ti,ab OR chile:ti,ab OR ‘costa rica’:ti,ab OR china:ti,ab OR kazakhstan:ti,ab OR ‘russian federation’:ti,ab OR russia:ti,ab OR vietnam:ti,ab OR estonia:ti,ab OR colombia:ti,ab OR portugal:ti,ab OR ecuador:ti,ab OR albania:ti,ab OR guadalupe:ti,ab OR turkiye:ti,ab OR lithuania:ti,ab OR lao:ti,ab OR ‘sao tomé e principe’:ti,ab OR afghanistan:ti,ab OR martinique:ti,ab OR brunei:ti,ab OR zimbabwe:ti,ab OR uzbekistan:ti,ab) AND [embase]/lim NOT ([embase]/lim AND [medline]/lim) AND ('article'/it)

Draft of search strategy in Web Of Science:

ALL=((Stomach OR Gastric*) AND (Neoplas* OR Cancer OR Tumor* OR Tumour* OR Carcinoma* OR Adenocarcinoma* OR Malignan*) AND (Endoscop* OR Gastroscop*) AND ("Mass Screening" OR Screening*) AND ("Intermediate Risk" OR "Moderate Risk" OR "Intermediate Incidence" OR "Moderate Incidence" OR West* OR Europ* OR tajikistan OR iran OR azerbaijan OR kyrgyzstan OR bhutan OR belarus OR peru OR mali OR chile OR "costa rica" OR china OR kazakhstan OR "russian federation" OR russia OR vietnam OR estonia OR colombia OR portugal OR ecuador OR albania OR guadalupe OR turkiye OR lithuania OR lao OR "sao tomé e principe" OR afghanistan OR martinique OR brunei OR zimbabwe OR uzbekistan))
