## Supplementary material 2 for "Effectiveness of Gastric Cancer Endoscopic Screening in Intermediate-Risk Countries – a Systematic Review and Meta-Analysis"

| **Variable** | **Definition / domain** |
| --- | --- |
| Endoscopic screening | Number of individuals screened by endoscopy |
| Frequency of screening | Frequency of endoscopic screening in years |
| Age range covered by screening | Age at start and end of screening |
| Screening adherence rate | % of invited individuals that did the screening |
| Number of biopsies | total number of biopsies performed in the study |
| Helicobacter Pylori diagnosis rate | % of individual screened that were diagnosed with *Helicobacter pylori* |
| Detection rate of pre-malignant lesions | Number of premalignant lesions (atrophic gastritis, intestinal metaplasia and low-grade intraepithelial neoplasia - formerly low-grade dysplasia) / Total number of upper endoscopies performed. |
| Gastric cancer detection rate | Number of gastric cancers diagnosed by screening endoscopy / Total number of screening upper endoscopies performed |
| Early gastric cancer detection rate | Number of early gastric cancers (high-grade intraepithelial neoplasia; mucosal adenocarcinoma) detected by endoscopic screening / Total number of gastric cancers detected by endoscopic screening |
| Lethality rate | Number of people who died from gastric cancer diagnosed by endoscopic screening / population at risk during the study period |
| 5-year survival rate in patients diagnosed with gastric cancer at screening | Percentage of patients diagnosed with stomach cancer at screening who live at least 5 years after diagnosis |
| Incremental Cost Effectiveness Ratio | Value (in euro) of implementing the upper endoscopy screening programme (or adding it to other existing screening programmes, e.g. endoscopic screening for colorectal cancer) |

Table 1 Variables related to outcomes of screening and cost-efficiency

Table 2. Other variables

| **Variable** | **Domain** |
| --- | --- |
| Study Bibliographic Reference | Reference |
| Country or countries of origin of the study | Country(ies) |
| Study design | Randomized controlled trial  Non-randomized controlled trial  Cohort  Case-control  Cross-sectional  Cost-effectiveness |
| Population /sample | Base population potentially to be screened by upper endoscopy |
| Participants | Age  Sex distribution |
