## Supplementary material 8 for "Effectiveness of Gastric Cancer Endoscopic Screening in Intermediate-Risk Countries – a Systematic Review and Meta-Analysis"

### Resultados

#### Effect Sizes and (Sampling Variances or Standard Errors)

Random-Effects Model (k = 22)

| | Estimate | se | Z | p | CI Lower Bound | CI Upper Bound |
|---|---|---|---|---|---|---|
| Intercept | 0.280 | 0.0547 | 5.13 | <.001 | 0.173 | 0.387 |
| | . | . | . | . | . | . |

*Nota.* Tau² Estimator: Restricted Maximum-Likelihood

**[3]**

Heterogeneity Statistics

| Tau | Tau² | I² | H² | R² | df | Q | p |
|---|---|---|---|---|---|---|---|
| 0.256 | 0.0657 (SE= 0.0203 ) | 100% | 43509.772 | . | 21.000 | 74943.348 | <.001 |

##### Forest Plot

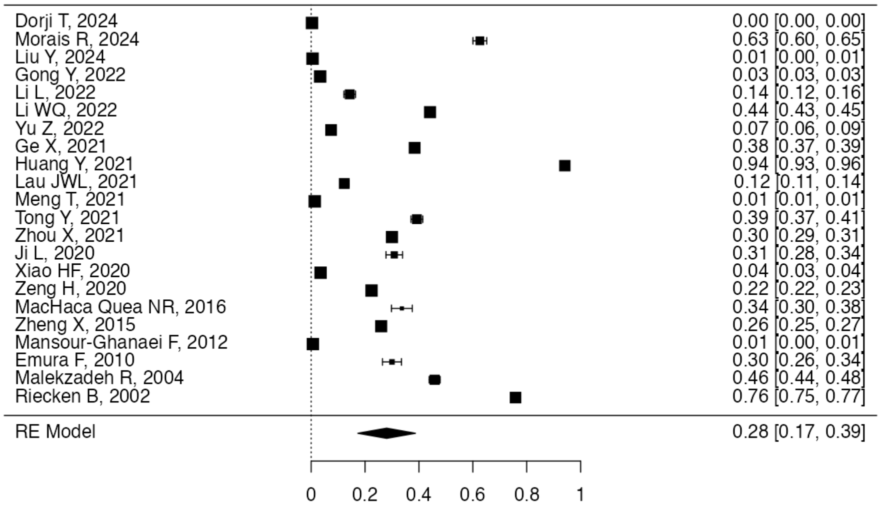


**[3]**

Publication Bias Assessment

| Test Name | value | p |
|---|---|---|
| Fail-Safe N | 380415.000 | <.001 |
| Kendalls Tau | 0.195 | 0.217 |
| Egger's Regression | 1.972 | 0.049 |

*Nota.* Fail-safe N Calculation Using the Rosenthal Approach

##### Funnel Plot

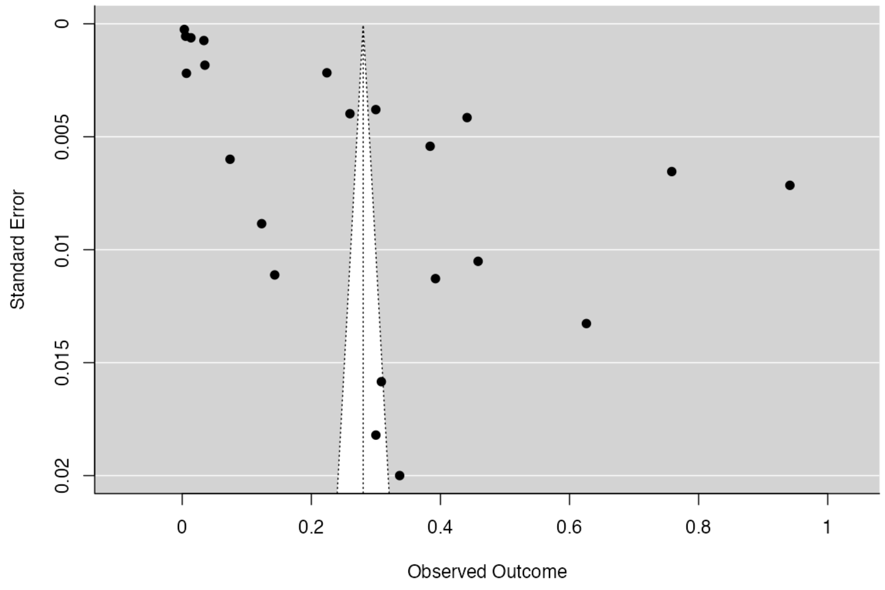


**[3]**

Two One-Sided Tests Equivalence Testing

| Z-Value Lower Bound | P-Value Lower Bound | Z-Value Upper Bound | P-Value Upper Bound | LL_CI_TOST | UL_CI_TOST | LL_CI_ZTEST | UL_CI_ZTEST |
|---|---|---|---|---|---|---|---|
| 14.272 | <.001 | -4.019 | 0.000 | 0.190 | 0.370 | 0.173 | 0.387 |

**[4]**

#### Two One-Sided Tests Equivalence Testing: Text Summary

```
The equivalence test was significant, Z = -4.019, p = 0.0000293, given equivalence bounds of -0.500 and 0.500 and an alpha of 0.05.
The null hypothesis test was significant, Z = 5.127, p = 0.000000295, given an alpha of 0.05.
NA
```

#### Equivalence Test Plot

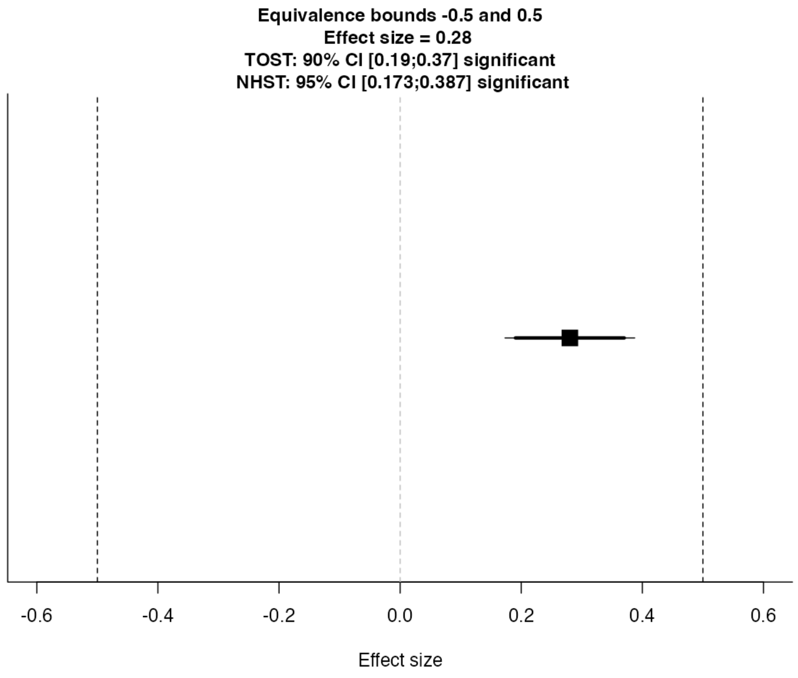


**[4]**

### Referências


**[1]** The jamovi project (2023). *jamovi*. (Version 2.4) [Computer Software]. Retrieved from https://www.jamovi.org.

**[2]** R Core Team (2022). *R: A Language and environment for statistical computing.* (Version 4.1) [Computer software]. Retrieved from https://cran.r-project.org. (R packages retrieved from CRAN snapshot 2023-04-07).

**[3]** Viechtbauer, W. (2010). Conducting meta-analyses in R with the metafor package. *Journal of Statistical Software. link, 36*, 1-48.

**[4]** Lakens, D. (2017). Equivalence tests: A practical primer for t-tests, correlations, and meta-analyses. *Social Psychological and Personality Science. link, 1*, 1-8.
