## Supplementary material 10 for "Effectiveness of Gastric Cancer Endoscopic Screening in Intermediate-Risk Countries – a Systematic Review and Meta-Analysis"

### Resultados

#### Effect Sizes and (Sampling Variances or Standard Errors)

Random-Effects Model (k = 15)

| | Estimate | se | Z | p | CI Lower Bound | CI Upper Bound |
|---|---|---|---|---|---|---|
| Intercept | 0.736 | 0.0660 | 11.2 | < .001 | 0.607 | 0.866 |
| | . | . | . | . | . | . |

*Nota.* Tau² Estimator: Restricted Maximum-Likelihood

**[3]**

Heterogeneity Statistics

| Tau | Tau² | I² | H² | R² | df | Q | p |
|---|---|---|---|---|---|---|---|
| 0.245 | 0.0598 (SE= 0.0244 ) | 100% | 5313659.093 | . | 14.000 | 1609.038 | < .001 |

#### Forest Plot

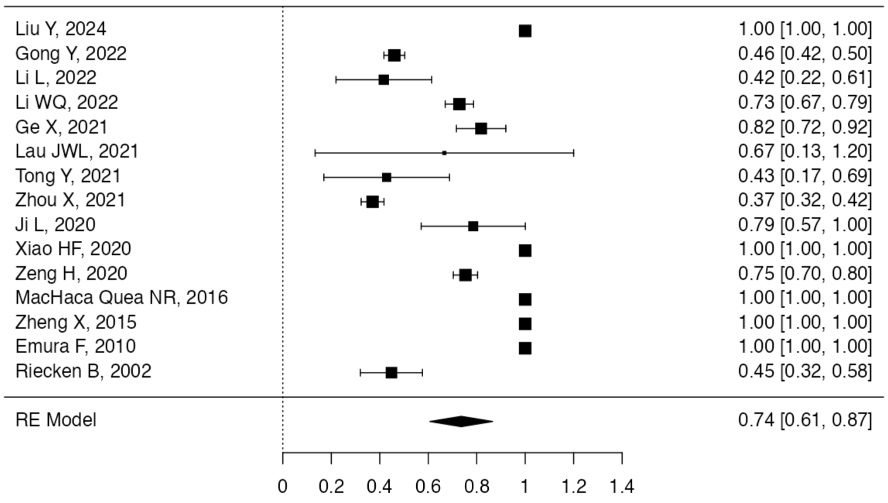


**[3]**

Publication Bias Assessment

| Test Name | value | p |
|---|---|---|
| Fail-Safe N | 2451864682.000 | < .001 |
| Kendalls Tau | -0.238 | 0.239 |
| Egger's Regression | -1.955 | 0.051 |

*Nota.* Fail-safe N Calculation Using the Rosenthal Approach

#### Funnel Plot

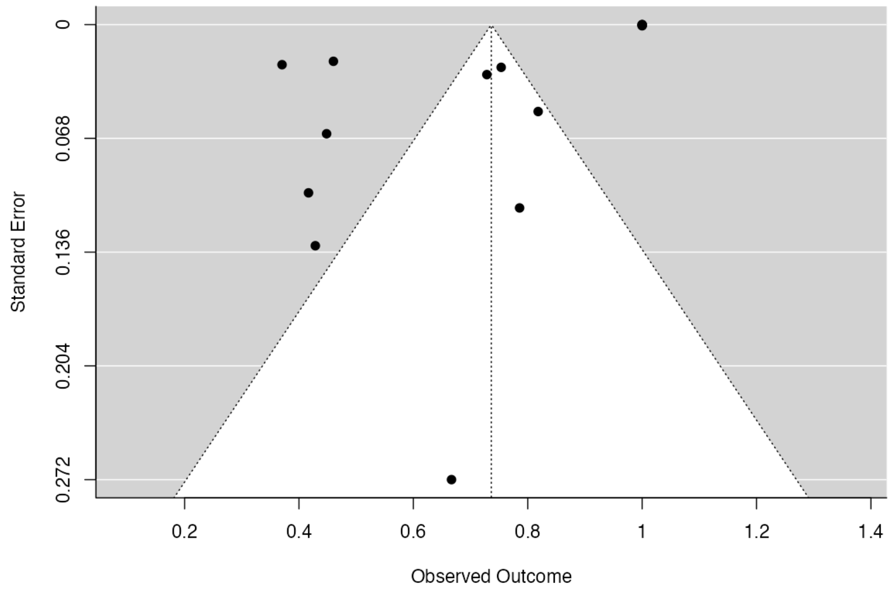


[3]

Two One-Sided Tests Equivalence Testing

| Z-Value Lower Bound | P-Value Lower Bound | Z-Value Upper Bound | P-Value Upper Bound | LL_CI_TOST | UL_CI_TOST | LL_CI_ZTEST | UL_CI_ZTEST |
|---|---|---|---|---|---|---|---|
| 18.744 | <.001 | 3.582 | 1.000 | 0.628 | 0.845 | 0.607 | 0.866 |

[4]

#### Two One-Sided Tests Equivalence Testing: Text Summary

```
The equivalence test was non-significant, Z = 3.582, p = 1.000, given equivalence bounds of -0.500 and 0.500 and an alpha of 0.05.
The null hypothesis test was significant, Z = 11.163, p = 0.0000000000000000000000000000617, given an alpha of 0.05.
NA
```

#### Equivalence Test Plot

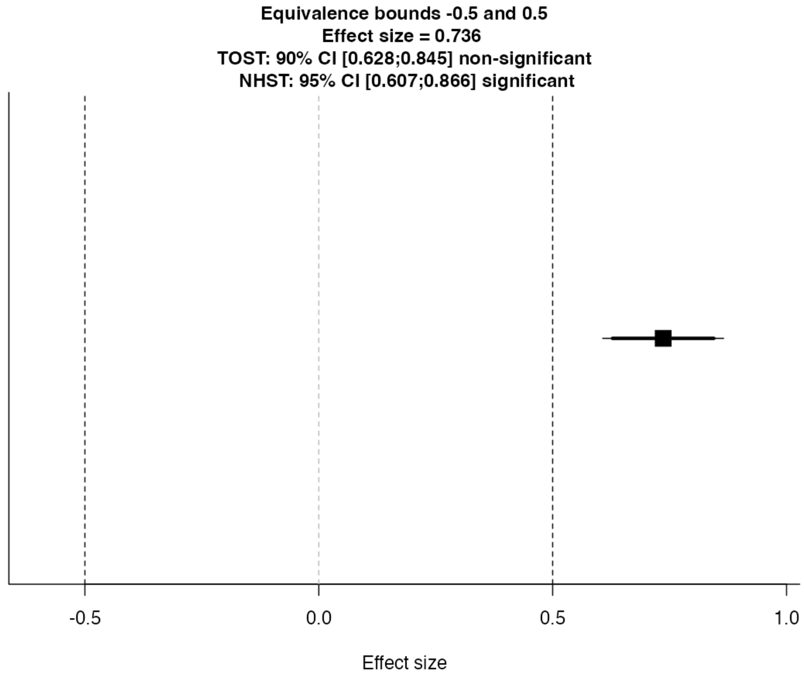


[4]

#### Referências

**[1]** The jamovi project (2023). *jamovi*. (Version 2.4) [Computer Software]. Retrieved from https://www.jamovi.org.

**[2]** R Core Team (2022). *R: A Language and environment for statistical computing.* (Version 4.1) [Computer software]. Retrieved from https://cran.r-project.org. (R packages retrieved from CRAN snapshot 2023-04-07).

**[3]** Viechtbauer, W. (2010). Conducting meta-analyses in R with the metafor package. *Journal of Statistical Software.* *link*, *36*, 1-48.

**[4]** Lakens, D. (2017). Equivalence tests: A practical primer for t-tests, correlations, and meta-analyses. *Social Psychological and Personality Science.* *link*, *1*, 1-8.
